# Natural and cryptic peptides dominate the immunopeptidome of atypical teratoid rhabdoid tumors

**DOI:** 10.1101/2021.06.11.21258679

**Authors:** Ana Marcu, Andreas Schlosser, Anne Keupp, Nico Trautwein, Pascal Johann, Matthias Wölfl, Johanna Lager, Camelia Monoranu, Juliane Sarah Walz, Lisa M. Henkel, Jürgen Krauß, Martin Ebinger, Martin Schuhmann, Ulrich Thomale, Torsten Pietsch, Erdwine Klinker, Paul G. Schlegel, Florian Oyen, Yair Reisner, Hans-Georg Rammensee, Matthias Eyrich

## Abstract

Atypical teratoid/rhabdoid tumors (AT/RT) are highly aggressive CNS-tumors of infancy and early childhood. Hallmark is the surprisingly simple genome with inactivating mutations or deletions in the SMARCB1 gene as the oncogenic driver. Nevertheless, AT/RTs are infiltrated by immune cells and even clonally expanded T cells. However, it is unclear, which epitopes T-cells might recognize on AT/RT cells. Here, we report a comprehensive MS-based analysis of naturally presented HLA-class-I and class-II ligands on 23 AT/RTs. Comparative HLA ligandome analysis of the HLA-ligandome revealed 55 class-I and 139 class-II tumor-exclusive peptides. No peptide originated from the SMARCB1-region. In addition, 61 HLA-class I tumor-exclusive peptide sequences derived from non-canonically translated proteins. Combination of peptides from natural and cryptic class I and class II origin gave optimal representation of tumor cell compartments. Substantial overlap existed with the cryptic immunopeptidome of glioblastomas but no concordance was found with extracranial tumors. More than 80% of AT/RT-exclusive peptides were able to successfully prime CD8^+^ T-cells, whereas naturally occurring memory responses in AT/RT-patients could only be detected for class-II epitopes. Interestingly, >50% of AT/RT-exclusive class-II ligands were also recognized by T-cells from glioblastoma patients but not from healthy donors. These findings highlight that AT/RTs, potentially paradigmatic for other pediatric tumors with a low mutational load, present a variety of highly immunogenic HLA-class-I and class-II peptides from canonical as well as non-canonical protein sources. Inclusion of such cryptic peptides into therapeutic vaccines would enable an optimized mapping of the tumor cell surface, thereby reducing the likelihood of immune evasion.

**One Sentence Summary:** The HLA-ligandome of atpyical teratoid-rhabdoid tumors contains immunogenic, tumor-exclusive peptides derived from natural and cryptic source proteins.

## Introduction

Atypical teratoid/rhabdoid tumors (AT/RTs) are malignant tumors of infancy and early childhood. They represent 50% of brain tumors in children under one year. Despite multimodal therapy, consisting of neurosurgery, polychemotherapy, and, whenever possible in these small children, radiotherapy, 5-year event-free survival is only 30% (*1*). One hallmark of this cancer are the surprisingly simple genomes, the oncogenic driver is usually restricted to a homozoygous deletional mutation in the SMARCB1 gene (*2*). More recently, AT/RTs have been categorized into the TYR-, SHH-, and MYC-subgroups which display distinct molecular and clinical features (*3, 4*). In cancer mutation rankings, AT/RTs are frequently considered the human cancer with the lowest mutational load (*5*). Although tumor mutational burden has emerged as an important biomarker for response to immunotherapy in certain adult cancer (*6*), this parameter is far from being exclusive. Immunohistological studies have revealed that AT/RTs can contain considerable infiltrations of immune cells including T cells (*7*). More recent RNA-seq data suggested that in the SHH-and MYC-subgroups of AT/RTs macrophages play an important role in tumor cell progression (*8*) and that these two subgroups also contain gene signatures of clonally expanded CD4^+^ and CD8^+^ T cells (*9, 10*). Attempts to categorize immune cell infiltrates by DNA methylation data identified three immune microenvironment clusters, but no clear difference between immune cells in AT/RT subtypes (*11*). Finally, tumor-specific CD8^+^ T-cell responses were reported in a small case series of AT/RT-patients receiving tumor-specific therapeutic vaccines (*12*). These data provide indications that from an immunological perspective, this tumor entity may not be entirely cold. However, the targets of a possible T-cell recognition are unknown so far. To address this issue, we performed a comprehensive analysis of AT/RT-specific T-cell epitope candidates. Due to the extremely low mutational load in AT/RTs we considered neoantigens derived from non-synonymous mutations as an unlikely source of candidate peptides and therefore focused our investigation on peptides eluted from HLA-class I and II molecules of primary tumor tissue. Our intention was to develop a strategy for vaccine design in low mutational tumors in which a maximal representation of the entire ligandome of the tumor cell is given, thereby preventing immune escape. Thus, our mass spectrometry (MS)-based discovery workflow and downstream computational analysis took into account not only tumor-associated antigens derived from canonical non-mutated or overexpressed natural proteins but also antigens from non-canonical or cryptic sequences, which have recently been reported to account for up to 15% of the immunopeptidome (*13, 14*). Our data demonstrate that AT/RTs express a plethora of tumor-specific and immunogenic HLA class I and class II ligands from highly diverse origin. Cryptic peptides represent a hitherto untapped source of potential T-cell epitopes which should be considered for personalized tumor vaccine designs.

## Results

### Immunohistochemical landscape of AT/RT tissue

First, we analyzed an independent cohort of n=17 AT/RT tissues for the prevalence of T cells, HLA and PD-L1 molecules. Diagnosis of AT/RT was confirmed by central neuropathological review in all cases. 1.8 ± 5.1 % of all cells within AT/RT tissues represented CD3^+^ T cells. HLA class I and to a lesser degree also class II molecules were found to be expressed on tumor cells. PD-L1 expression level on tumor cells was 5.1 ± 6.8% as determined by the Cologne scoring system (Fig. 1 A+B). Linear regression revealed a weak inverse correlation between HLA class I and CD3 expression (Fig. 1C) and positive correlation between HLA class I and PD-L1 (Fig. 1D). From one of the freshly obtained tumor samples we were able to create a primary rhabdoid cell line. On this cell line we could confirm constitutive expression of HLA class I and IFNγ-inducible upregulation of HLA class II and PD-L1 molecules (Fig. 1E).

**Fig. 1.**
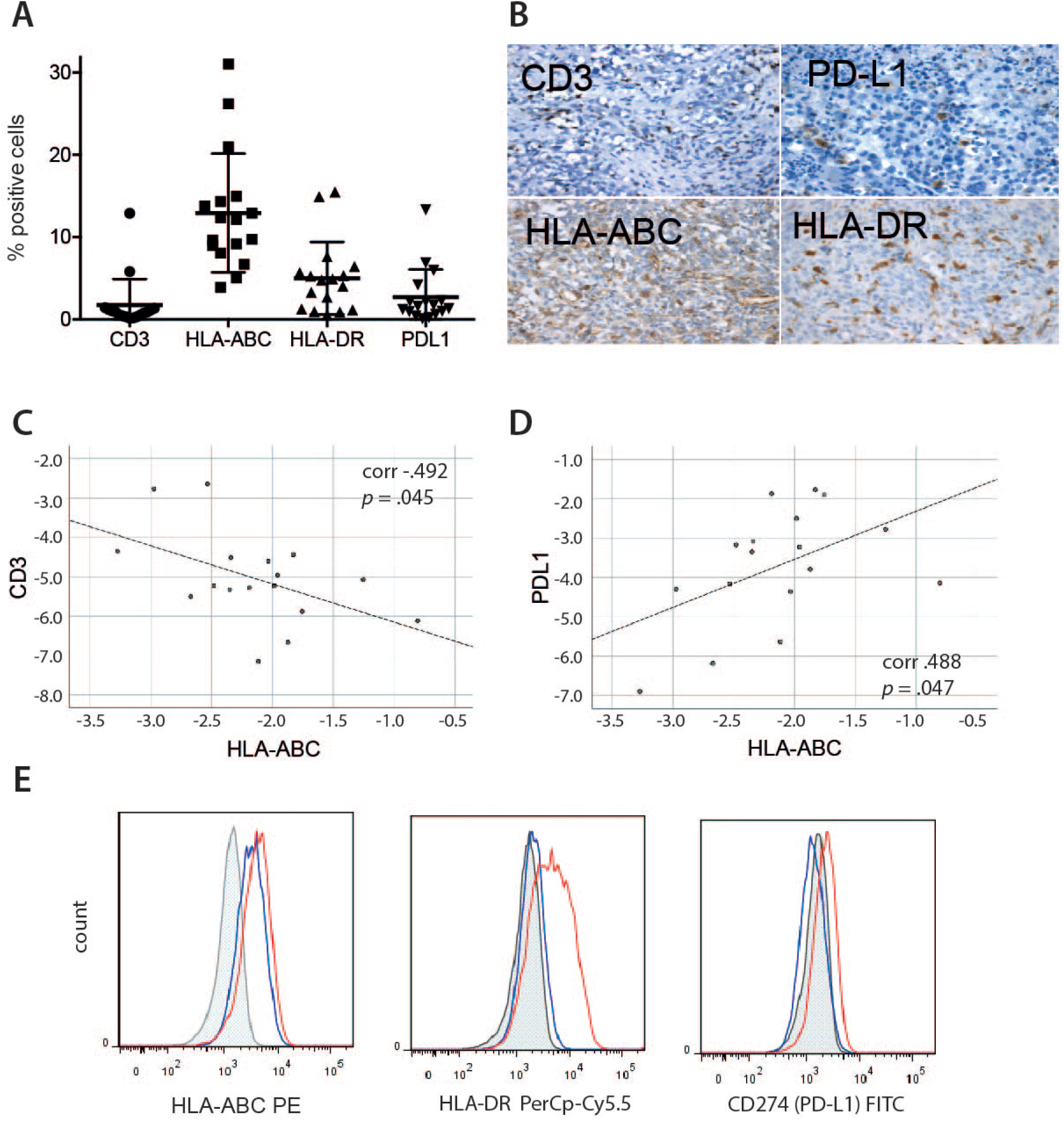
Immunohistochemistry of AT/RT tissues. n=17 AT/RT tissues were evaluated for expression of CD3, HLA-ABC, HLA-DR, and PD-L1 on tumor cells. **(A)** Mean values ± standard deviations for all four markers, 10 visual fields were evaluated in each tumor sample. **(B)** Representative examples for each of the four markers. **(C and D)** Linear regression analysis of HLA-ABC expression against CD3^+^ T-cell infiltration **(C)** and PD-L1 expression **(D). (E)** Flow cytometric analysis of one primary rhabdoid cell line. Shaded line represents unstained control, blue line constitutive expression, red line expression after IFNγ stimulation of tumor cell line.

### LC-MS/MS-based immunopeptidome profiling reveals AT/RT-associated HLA source proteins and HLA class I and II ligands eligible for immunotherapy approaches

23 AT/RT tumor samples could be retrieved from 5 tumor cell banks throughout Germany with a centrally confirmed diagnosis of an AT/RT. The cohort included a total of 51 different HLA class I allotypes, HLA-A*03:01 (n = 9) being the most frequent, followed by HLA-A*02:01 (n = 6), HLA-C*04:01 (n = 6), and HLA-C*07:01 (n = 6, figure S2). Among the world’s population, 99.9% of individuals carry at least one HLA class I allotype that is represented within the here analyzed AT/RT cohort (figure S2) (*15, 16*). The comparative cohort of benign tissue donors showed an HLA allotype population coverage of 99.8% (figure S2) and matched 100% of HLA-A allotypes (13/13), 72% of HLA-B allotypes (13/18), and 92% of HLA-C allotypes (11/12) of the AT/RT cohort.

LC-MS/MS analysis of these 23 AT/RT tumor samples identified 7,628 unique HLA class I peptides (range 13-1,372, mean 459 per sample) (Figure 2A) from 4,807 source proteins at a local PSM-level FDR of 5%. The in-house HLA class I benign cohort contained 282 tissues covering among others brain (n = 5) and cerebellum (n = 5) (HLA Ligand Atlas Paper Version 2017, extended with ligandomes from disease-free ovary). Based on these two datasets, we performed comparative HLA class I ligandome profiling to identify AT/RT-associated source proteins (Figure 2 B). We identified 55 HLA class I source proteins exclusively presented on AT/RT samples and from these 56 distinct HLA class I peptide ligands as candidates for downstream immunogenicity analysis (Figure 2B, table S1). The majority of AT/RT-associated source proteins (48/55) were individual to each AT/RT tumor, while only six were shared between two patients, and one was shared between tumor and ascites sample of the same patient.

**Fig. 2.**
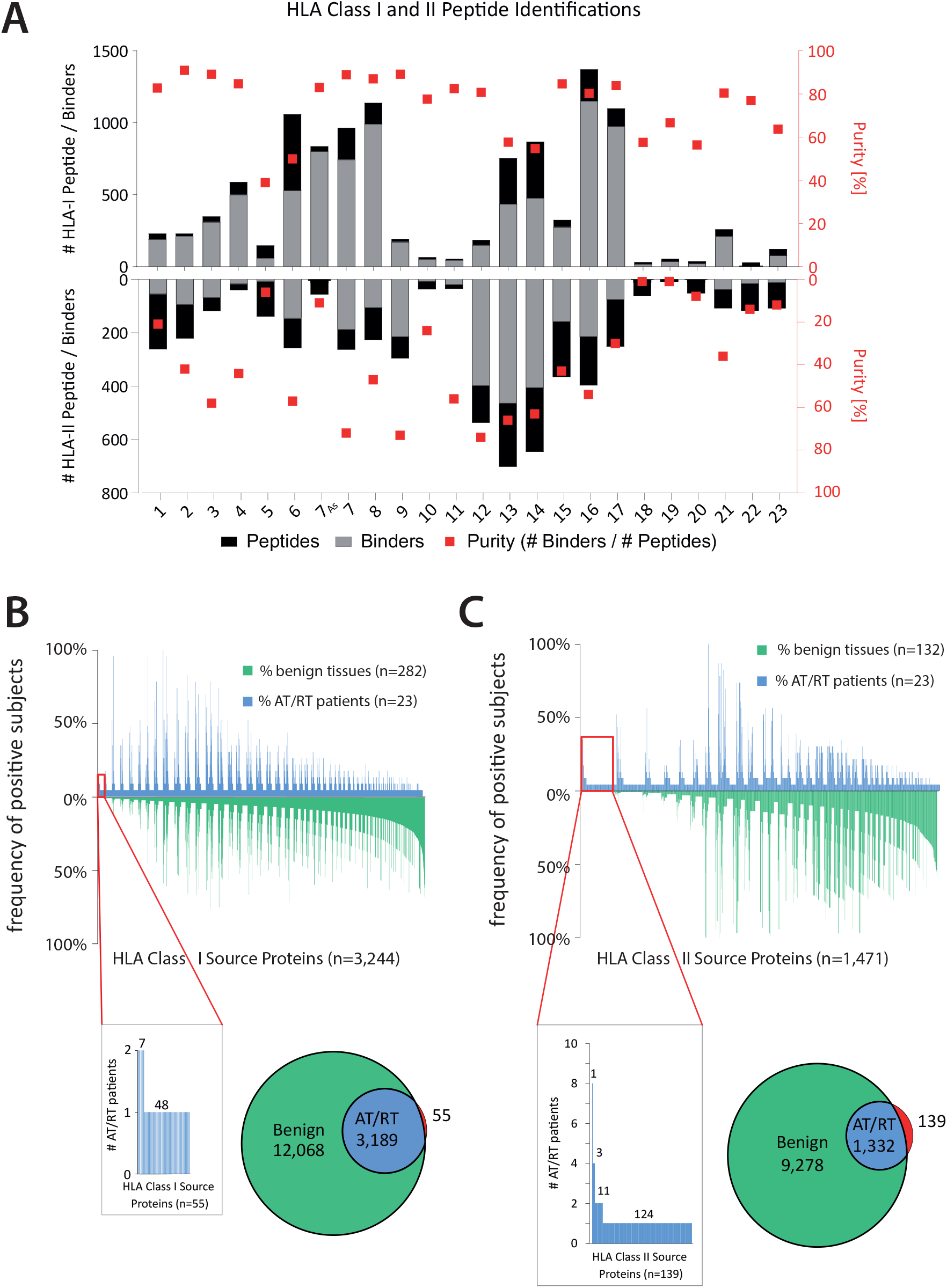
HLA Class I and II Peptide Identifications. **(A)** Number of identified peptides in each tumor sample for HLA class I (upward columns) and class II (downward colums). Grey *vs*. black shading illustrates predicted or non-predicted binding of peptides to self-HLA allotypes in the respective patients. Red squares (right y-axis) show purity of peptide pools, i.e. the ratio of predicted binding vs. total number of identified peptides. **(B)** Waterfall plot demonstrating overall source protein identifications for HLA class I ligands (x-axis) in benign (green, downward columns) and AT/RT tissue (blue, upward columns) and their frequency in the respective patient cohort. Tumor-exclusive are only the source proteins in the far left red boxes which have no corresponding counterpart in any benign tissue (red parts of Venn diagrams). **(C)** Same illustration for HLA class II source proteins.

With respect to HLA class II, 30 distinct class II allotypes were identified in the AT/RT patient cohort, with HLA-DQB1*03:01 (n = 27) being the most frequent, followed by HLA-DRB1*03:01 (n = 20) and HLA-DRB1*15:01 (n = 20). Among the world’s population, 99,7% of individuals carry at least one HLA class II allotype encompassed in the AT/RT cohort, with an average hit number of 3.71 per individual (figure S2).

LC-MS/MS analysis of the AT/RT HLA ligandome revealed 3,827 distinct HLA class II peptide sequences (range: 9 – 698, mean per sample 214) (Figure 2A) originating from 1,517 source proteins at 5% local PSM-level FDR. Our HLA class II benign cohort entails 132 ligandomes from benign samples, covering among other tissues, 5 cerebellum and 5 brain samples. Unfortunately, most benign HLA class II ligandomes lack HLA class II typing. By comparing the AT/RT and benign tissue cohort regarding single mapped source proteins, we were able to identify 139 AT/RT-associated HLA class II source proteins associated to 122 distinct HLA class II ligands (Figure 2C, table S1). Remarkably, multiple AT/RT-associated source proteins were shared among patients: one was presented in 8 different AT/RT samples, three additional source proteins were shared between four AT/RT patients and 11 were shared between two patients. As observed for HLA class I AT/RT-associated source proteins, the majority (124) was unique to each patient (Figure 2C).

### Cryptic peptides in AT/RTs ligandome

Next, we extended our search to identify cryptic HLA class-I peptides from allegedly non-coding regions of the human genome. To this end, we applied a recently introduced approach (Peptide-PRISM) that combines *de novo* peptide sequencing, highly efficient string search, mixture modeling, and search space stratified filtering of the false discovery rate (FDR) (*13*). *De novo* sequencing was performed for all LC-MS/MS data of the 23 AT/RT tumors samples with PEAKS X. Peptide-PRISM was used to match all *de novo* sequencing candidates against the 3-frame translated human transcriptome and the 6-frame translated human genome. 209 cryptic peptides were identified (FDR 10%), 158 of which were predicted as binder (rank < 2%) to one of the corresponding patient HLA alleles by NetMHCpan 4.0. The distribution of the 158 cryptic peptides among the different categories is shown in Fig. 3A. The highest number of peptides derived from non-canonical ORFs close to the canonical translation start site (categories UTR5 and OffFrame). These cryptic peptides presumably originate from translational misinitiation. An example of such a peptide is shown in Fig. 3B.

**Fig. 3.**
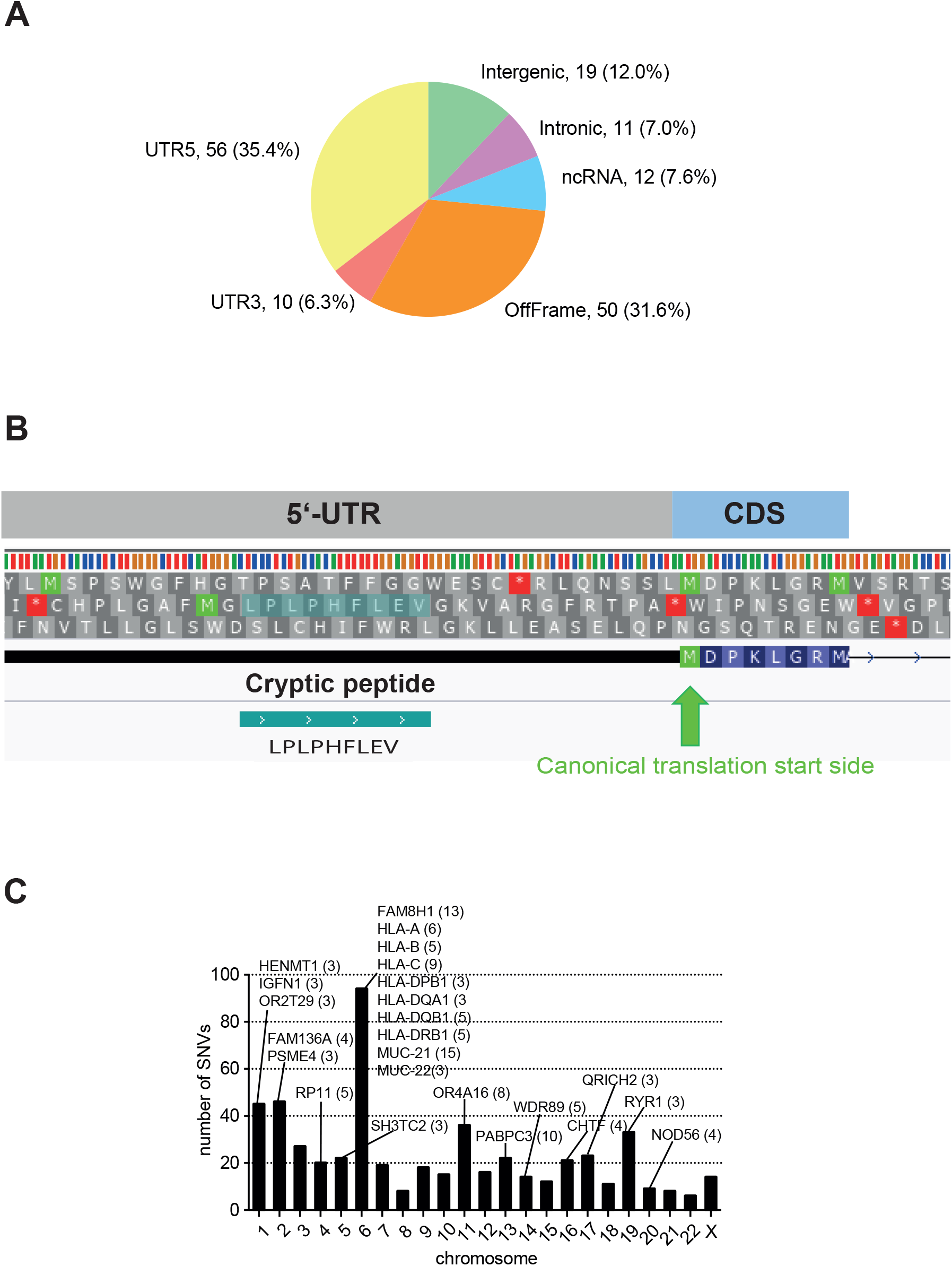
Identification of cryptic class I peptides. 158 class I peptides have been identified and predicted as binder to one of the corresponding patient alleles in AT/RT tumor samples. **(A)** Distribution of identified cryptic peptides among genomic categories. **(B)** Example of a cryptic HLA peptide from 5’-UTR. The peptide was identified in two AT/RT patients, but not in benign tissue. The peptide is predicted as a strong binder (rank < 0.5%) by NetMHCpan in both patients for HLA-B*51:01 and HLA-B*07:02, respectively. **(C)** Most frequently mutated proteins and their localization on the 23 chromosomes. Numbers in parentheses indicates the number of different mutations found in the respective protein.

In order to identify potentially tumor-associated or tumor-specific cryptic peptides, we filtered out all cryptic peptides that have previously been identified within the human HLA ligand atlas project (https://hla-ligand-atlas.org) (*17*). 61 of the 158 cryptic binding peptides were identified only in AT/RT tumor samples, but not in benign tissue (table S2). In addition, we compared the cryptic peptides identified in AT/RT samples with cryptic peptides previously identified in glioblastoma (GBM) tumor tissue, as well as plasma-soluble HLAs (*18*). A complete list of all cryptic peptides from these samples has been recently published (*13*). 20 of the 61 (33%) cryptic peptides have been recurrently identified in either at least two AT/RT patients (9/61, 15%) or AT/RT and GBM patients (11/61, 18%), e.g. the cryptic peptide LSLEEGIVEV derived from an intron of HIPK3 is identified recurrently in 5 AT/RT patients (table S2).

We selected six cryptic peptides from different categories for analyzing their immunogenicity (see below). Fragment ion spectra of these six synthetically preparated peptides were generated for validating the peptide identification. For all but one peptide (MPISLVQLL) comparison of the fragment ion spectra obtained from AT/RT samples with the fragment ion spectra of the corresponding synthetic peptide confirmed the initial peptide identification (figure S3).

### Nonsynonymous mutations in AT/RTs

Furthermore, we were interested to examine whether the oncogenic SMARCB1 mutation can give rise to immunogenic peptides. After ligandome analysis, additional tissue was left in 18 cases, in 9 of these also PBMCs were available. In the cases with tumor tissue only, MPLA sequencing of the SMARCB1 locus was performed. In these assays, only homo-or heterozygous deletions but no SNVs of the SMARCB1 locus were detectable, making the existence of neoepitopes in this region unlikely (table S3). In the other nine AT/RT cases we isolated genomic DNA from fresh-frozen AT/RT tissues and corresponding PBMCs for whole-exome sequencing (WES). As expected, the overall number of non-synonymous mutations was low (median 16, range 1 – 155). Five tumors harbored ≤ 7 SNV, whereas four tumors displayed a higher number of SNVs (99, 112, 137, and 155). Two of these belonged to the SHH-, one each to the TYR-and MYC-group, respectively. Also in WES, no non-synonymous mutations in the SMARCB1 locus could be identified. The 24 most frequently mutated genes and their chromosomal location are shown in Figure 3C.

To identify potential neoantigenic epitopes from these SNVs, overlapping 9-mer peptide libraries moving the mutation from pos 1-9 were generated and analyzed for predicted binding to all HLA alleles of the individual patient. The peptide with the best binding score for any of the patients’ HLA alleles was selected. A complete list of all *in silico* predicted neoantigens is provided in table S4. None of these neoantigenic peptide sequences could be verified in the MS data.

### Immunogenicity of ligandome peptides

To assess potential immunogenicity of all identified, AT/RT-associated HLA class I peptides, we first compared *in silico* predicted MHC binding affinity using NetMHCpan 4.0. Whereas binding affinity of neoantigenic and ligandome cryptic peptides was almost identical, ligandome-identified canonical class I peptides displayed a significantly higher binding affinity, suggesting that these peptides have undergone an *in vivo* selection process in the class I peptide loading complex (Fig. 4A).

**Fig. 4.**
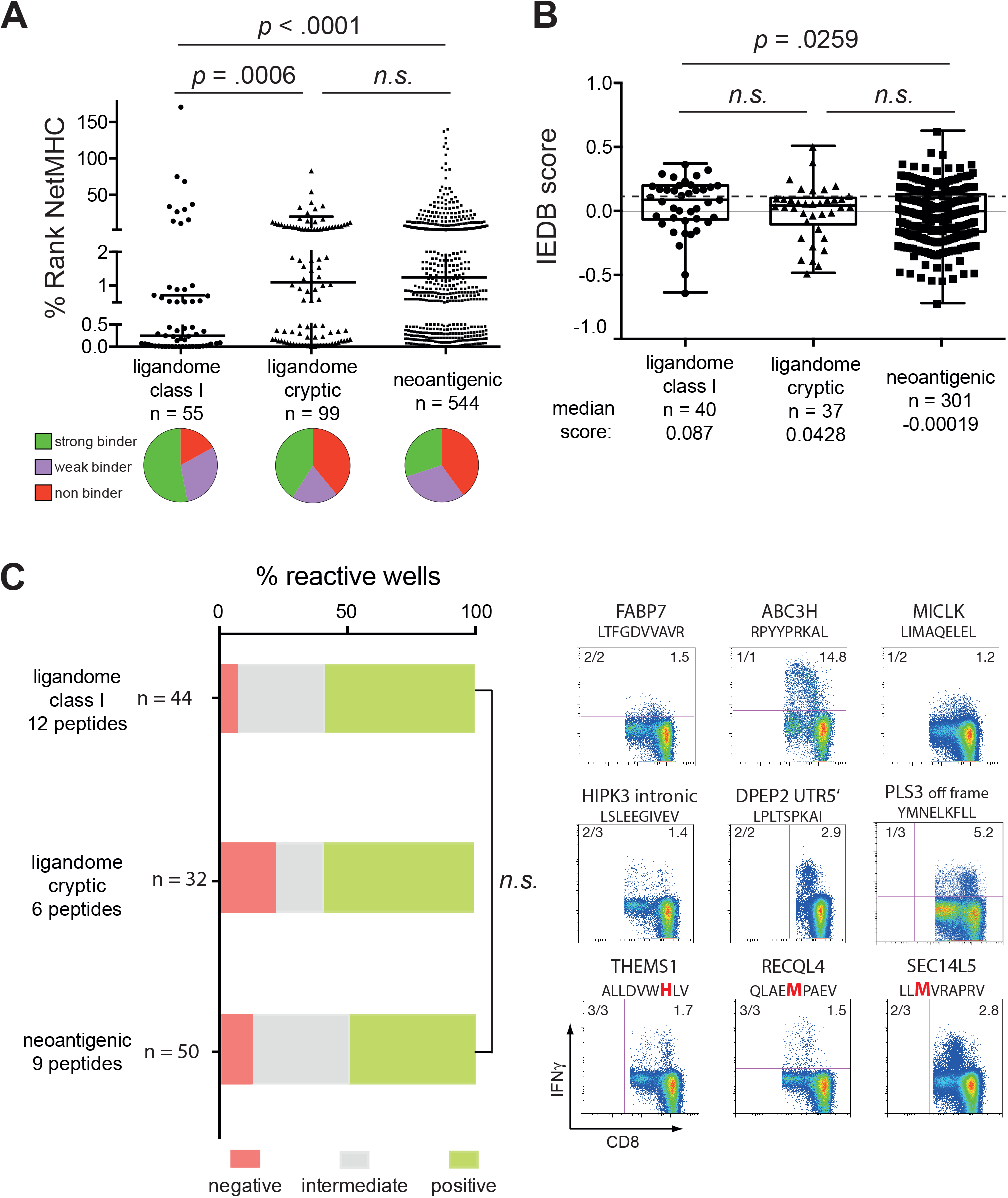
Immunogenicity of HLA class I peptides. **(A)** Peptides sequences of natural and non-canonical HLA-ligands identified by MS as well as *in silico* predicted neoepitopes were ranked according to the NetMHCpan4.0 algorithm. The best binding HLA allele from each tumor sample was chosen. Pie charts below the graph show classification of %rank values into strong (≤0.5), weak (0.5-2) and non-binders (>2), n indicates the total number of peptides per group. **(B)** Peptides with an HLA-ligand listed in the IEDB data base were alternatively scored for immunogenicity according to the IEDB alogrithm, n indicates the number of peptides which could be submitted to the IEDB server. **(C)** Highly purified naïve CD8^+^ T cells from 5 healthy HLA-A*02:01 (n=4) and -B07*02 (n=1) positive donors were cocultured for 12 days with peptide-loaded autologous DCs in the presence of IL-7, -15, and -21, restimulated and assayed for IFNγ/TNFα production. A response 3x above background (empty DCs) was considered as positive (green bars), responses above background but not positive were defined as intermediate (grey bars). N indicates the total number of wells tested in the respective peptide group. Pseudo-color dot plots on the right side display three representative examples of positive responses for ligandome natural and cryptic peptides and neoantigenic epitopes, respectively. For the complete data set of priming assays please refer to fig. S3. Numbers in the upper right quadrant show percentages of CD8^+^IFNγ^+^, numbers in the upper left quadrant indicate number of positive experiments against the amount of donors tested. The bold and red colored amino acid indicates the mutation. P-values were determined using Mann-Whitney or Kruskal-Wallis test.

MHC-I binding algorithms mainly rely on binding of certain amino acids to anchor residues at P1, P2, and P9 of the MHC-I peptide binding groove. Recently, several *in silico* immunogenicity prediction tools have used alternative computational methods to assess T-cell receptor recognition of a peptide MHC complex, e.g. based on the presence of large, aromatic residues at P4, P5, and P6 of a peptide (IEDB database) or the hydrophobicity, molecular size and polarity of whole protein sequences (VaxiJen server). The IEDB database requires knowledge of the individual peptide sequence and only a limited set of validated HLA-alleles can be analyzed with this tool. For IEDB, we considered only peptides with the validated length of 9 amino acids. IEBD generally yielded lower frequencies of peptides with a positive prediction (37.8 – 52.3%) than the VaxiJen platform (62.3 – 77.9%). Using the IEDB prediction algorithm AT/RT ligandome class I peptides had higher immunogenicity prediction scores than neoantigenic peptide sequences (Fig. 4B). In contrast, the VaxiJen server favored neoantigenic and ligandome cryptic over ligandome class I source proteins (data not shown). There was no correlation between MHC binding affinity and either IEBD or VaxiJen scores (data not shown).

In addition to these *in silico* predictions, we tested the immunogenicity of the identified peptides in an *in vitro* priming assay. This assay uses highly-purified naïve CD8^+^ T cells from healthy donors as starting population and autologous DCs as stimulators, thereby closely mimicking a vaccination scenario *in vivo* (*19, 20*). We focused on two HLA-alleles with a high prevalence in our general population (HLA-A*02:01 and -B*07:02) and tested all identified peptides for binding to these two alleles. From the resulting strong binders we choose 12, 6, and 9 peptides from the list of ligandome-identified canonical class I, cryptic, and neoantigenic peptide pools, respectively. 92% (11 out of 12), 83% (5 out of 6), and 89% (8 out of 9) of the tested peptides yielded at least one positive well with no significant difference between the groups (Fig. 4C, the complete data set of priming assays is shown in fig. S4), confirming that the majority of peptides from the different pools can be recognized by naïve T cells.

Furthermore, we investigated whether memory responses against these peptides can be detected in AT/RT patients. PBMCs from 4 AT/RT patients were assayed in restimulation experiments for IFNγ-response against peptides fitting their HLA restrictions. For HLA class I peptides we screened all peptides for potential binding to HLA-A and -B alleles from these four patients and identified 17, 18, 23, and 32 suitable peptides for restimulation experiments. As demonstrated in Fig. 5A only one ligandome peptide (FPILSTILL from the CCG4 source protein) showed a positive response in one patient whereas all other experiments remained negative, confirming that constitutive CD8^+^ T-cell responses against tumor-associated peptides without prior vaccination are exceedingly rare. The same assay was repeated for the 18 HLA class II peptides which were identified more than once in AT/RT tumor samples with gating on CD3^+^CD4^+^ T cells (Fig. 5B). Notably, 15/18 class II peptides showed at least one positive response, 2 peptides even gave positive results in three out of four patients tested. To analyze whether these responses are specific for patients with AT/RT, we repeated this experiment with PBMCs from three high-grade glioma (HGG) patients and three healthy controls. In HGG patients, 10/18 peptides yielded a positive response, however, only in one patient each. The LSIEEDVLAA peptide derived from THMS1 source protein (detectable in 8 different AT/RT samples) showed a more substantial response in one HGG patient (Fig. 5B). In PBMCs from healthy controls only 8 faint responses at the detection limit in singular patients were measurable. These data indicate that T_helper_ cells of AT/RT patients have been exposed to HLA class II ligandome peptides before which has resulted in memory formation in more than 80% of tested peptides. This phenomenon seems to be tumor-specific as such responses were less detectable in HGG patients and virtually absent in healthy donors. For HLA class I peptides no such memory formation could be observed.

**Fig. 5.**
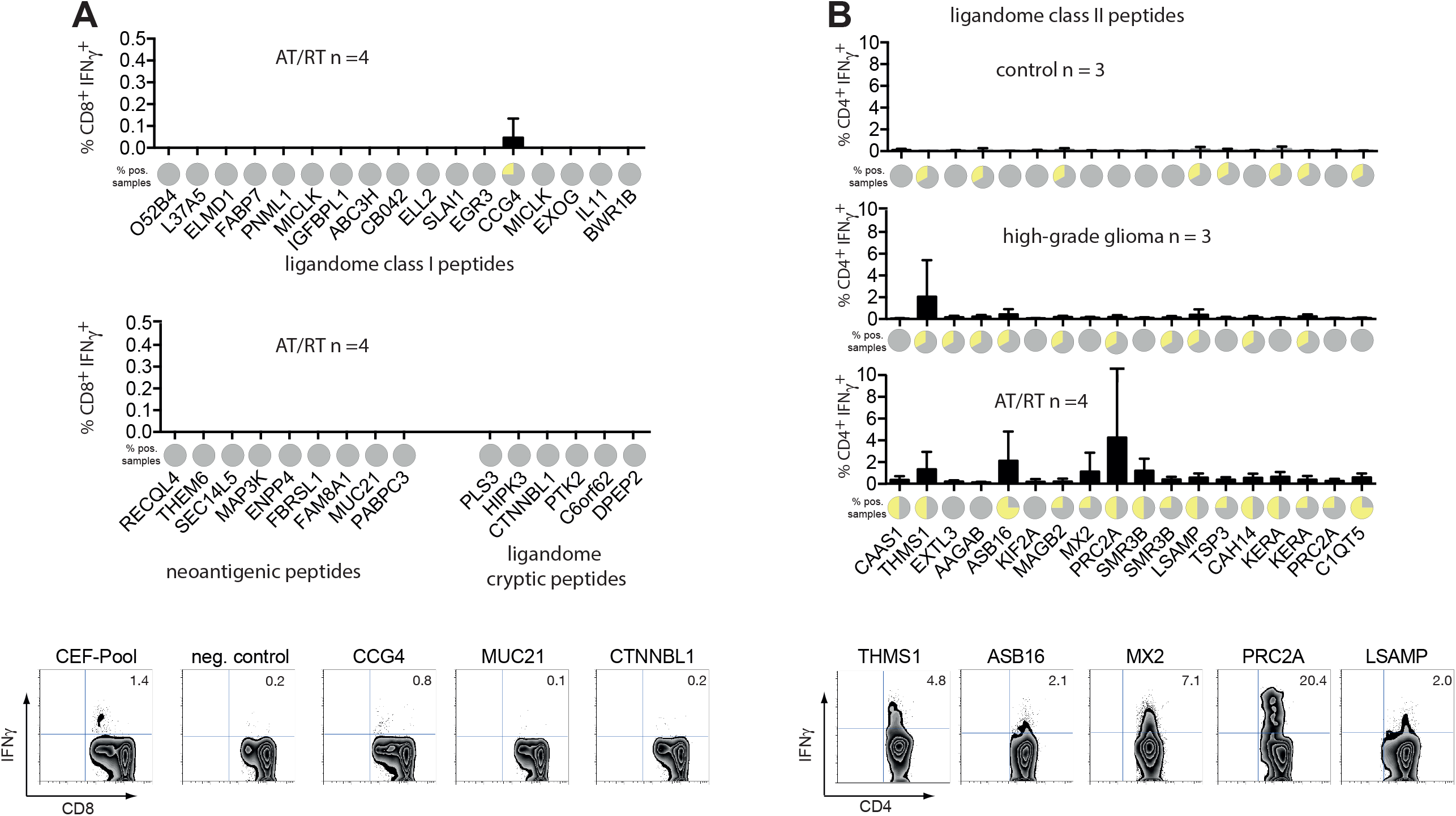
Recall responses against HLA class I and class II peptides in AT/RT patients. **(A)** Peptides sequences of natural and non-canonical HLA class I ligands identified by MS as well as *in silico* predicted neoepitopes were screened for binding to the HLA-A and -B alleles of 4 AT/RT patients, in whom PBMCs were available. The resulting 17, 18, 23, and 32 peptides were used to activate PBMCs from these 4 patients. Column graphs show frequencies of CD8^+^IFNγ^+^ T cells after 1-2 rounds of restimulation. Pie charts above source protein names display the frequency of positive tests (>3x above background, in yellow). Zebra plots below the charts show one representative example of the onliest positive result in a natural ligandome peptide and two negative wells from a cryptic and neoantigenic peptide from the same patient plus controls. **(B)** The 18 ligandome class II peptides which were detected in more than one tumor sample were used to activate CD4^+^ T cells from 4 AT/RT patients. PBMCs of three patients with high-grade gliomas and three healthy individuals served as controls. Column graphs show frequencies of CD4^+^IFNγ^+^ T cells after 1-2 rounds of restimulation. Pie charts above source protein names display the frequency of positive tests (>3x above background, in yellow). Zebra plots below the charts show five representative examples of positive responses to class II peptides from different AT/RT patients.

### Gene ontology and protein network analysis

Having identified tumor-exclusive peptides from four different sources (ligandome HLA class I and II peptides from natural proteins, ligandome class I cryptic, and neoantigenic peptides) we mined these data for existing overlap between the respective source proteins or pathway enrichment. Functional annotation using the DAVID Bioinformatics database revealed little similarity in terms of *biological process* or *molecular function* (table S5). Of note, the four different peptide sources covered distinct cellular compartments. In line with previous reports, HLA class I ligandome peptides were mainly derived from cytosolic proteins, whereas HLA class II ligandome source proteins were allocated to the plasma membrane. AT/RT neoantigens covered external and internal membrane proteins (plasma, Golgi, endo-/lysosomal membranes, junctions, MHC class II complex), whereas source proteins of ligandome cryptic peptides were primarily assigned to the nucleoplasm (Fig. 6A). Functional enrichment analysis of protein networks showed 4, 3, and 1 enriched protein domains within neoantigenic, cryptic and HLA class I source proteins, respectively, whereas for HLA class II no such enrichment could be detected (Fig. 6B). Likewise, only the network of mutated proteins displayed a significantly higher number of interactions than expected, with a cluster in HLA-and HLA-associated genes (Fig. 6C), which is consistent with the high number of mutations on chromosome 6 (Fig. 3C). Taken together, these data indicate that source proteins from different AT/RT-specific peptide pools cover a wide variety of biological functions, and show little enrichment of particular protein domains. However, at least in AT/RTs, the different peptide sources seem to emerge from distinct cellular compartments.

**Fig. 6.**
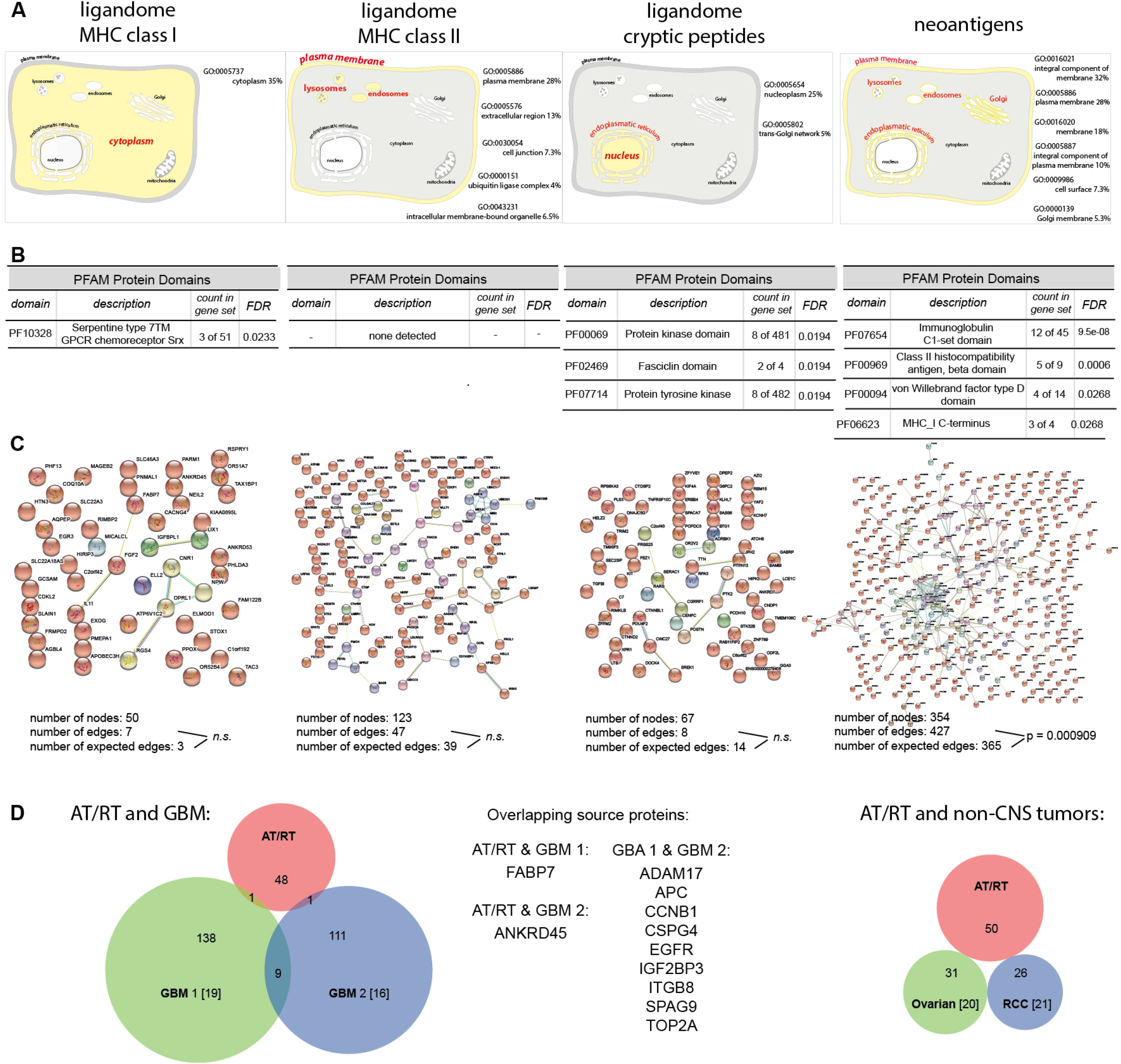
Gene ontology and network analysis of AT/RT peptides. (**A**) Functional annotation “cellular compartment” of the four different AT/RT peptides sources defined by DAVID Bioinformatics Resources 6.8. (**B**) Listing of enriched protein domains in the four source protein cohorts. (**C**) Protein network analysis using STRING. (**D**) Overlap of the AT/RT peptide pool with four published cancer immunopeptidome data sets: GBM 1 (*21*), GBM2 (*18*), ovarian cancer (*22*) and renal cell carcinoma (*23*).

### Overlap of AT/RT ligandome with other tumor-specific ligandomes

Finally, we matched our list of HLA class I AT/RT ligandome peptides from natural source proteins with published data sets of tumor-specific ligandomes from other cancer entities. Selection criterion for these comparators was their tumor-specificity, i.e. the exclusion of peptides also found in other benign tissues. We used two published data sets from glioblastoma (*18, 21*), and one from ovarian cancer (*22*) and renal cell carcinoma (*23*) each. While 6% of source proteins overlapped between the two GBM datasets, only 2% could be found simultaneously in AT/RT and GBM. No overlap existed between AT/RTs and the two non-CNS cancers (ovarian and renal, Fig. 6D). The more substantial overlap (33%) between AT/RT and GBM in the cryptic ligandome has been mentioned above.

## Discussion

Tumor immunosurveillance represents a difficult task for immunity, since cancer cells are a derivate of self and thymic selection has trained T-cells for self-tolerance. The vast majority of the several hundred thousand MHC molecules on a cancer cell is loaded with self-peptides. For tumor eradication both the affinity between peptide and MHC (*24*) and between TCR and peptide-MHC complex (*25*) has been demonstrated to be of critical importance. Therefore, the search for optimal anticancer T-cell epitopes focuses either on high affinity peptides from natural proteins, which are normally not expressed on benign tissue or on peptides derived from sequences, which are not part of the thymic Aire^+^ medullary epithelial cell library effective during T-cell selection. From the latter category, peptides covering a cancer-specific mutation have been intensely investigated in recent years. Although T-cell responses against such neoantigens derived from non-synonymous mutations undoubtedly exist and can be boosted by vaccination approaches (*26*), detection of neoantigenic peptides within the HLA-ligandome has been exceedingly rare (*27*). One study estimated that less than 1% of all non-silent mutations are represented in the HLA-ligandome (*28*). Given the fact that most tumors harbor less than 100 somatic mutations and that at least three identical neighboring peptide-MHC complexes are required to trigger cytotoxicity (*29*), the search for neoepitopes has been extended to so-called non-mutated neoantigens, i.e. epitopes derived from non-canonical translated or novel protein isoforms (*30*). This new category of HLA-bound epitopes has originally been discovered in the ligandome of normal cells, but recently also confirmed in adult tumors (*31, 32*). Pediatric cancers generally rank amongst tumors with a low mutational burden. AT/RTs are a typical representative of this category, as one deleting mutation in the SMARCB1 gene is sufficient for oncogenesis (*33*). Since SMARCB1 deletion leads to widespread epigenetic dysregulation, AT/RTs are considered epigenetic diseases. Yet, AT/RTs are infiltrated by immune cells (*7*), and subgroups of AT/RTs, especially the –MYC and –SHH subsets (*9, 11*), even show a preference for cytotoxic T cells (*9, 10*). Our IHC data confirm that approx. 2% of all cells within the AT/RT microenvironment are T cells. The negative correlation between CD3 and HLA-ABC could be interpreted as HLA-downmodulation as part of a reversible, epigenetically regulated tumor escape mechanism (*34*). However, the accumulation of mutations on chromosome 6 and the identification of MHC mutations as the only significant network cluster in gene ontology analysis of WES data could also point towards partial HLA-loss as an AT/RT-intrinsic ‘hard’ lesion (*35*). This issue, including strategies to restore HLA-expression, warrants further investigation. Furthermore, our data confirm that neoepitopes derived from non-synonymous mutations are not presented on the surface of AT/RT cells (at least not within our range of detection), as no SMARCB1-derived peptide could be identified. All analyzed patients in our cohort exhibited truncating mutations, making generation of mutation-derived peptides even more unlikely. Unexpectedly, 4 AT/RTs showed 100 or more mutations with no preference for one of the recently consented subgroups (*3, 4*), but also in these cases no neoantigenic peptide could be confirmed in the ligandome data.

Ligandome analyses have been performed in several adult cancer types such as melanoma (*27*), glioblastoma (GBM) (*21*), colorectal (*36*), lung (*37*), renal cell (RCC) (*38*), or ovarian cancer (*22*) as well as in hematological malignancies such as multiple myeloma (*39*) and leukemias (*40*). Our study in AT/RTs represents the first pediatric tumor entity analyzed. As in their adult counterparts, the AT/RT ligandome is primarily composed of peptide ligands from a wide variety of natural proteins with little overlap between individual samples. Overlap was higher for HLA class II peptides, as epitopes derived from THEMIS, AAGAB, ASB16, and EXTL3 proteins were found in more than two tumors in our cohort. These targets can be regarded as *bona fide* AT/RT HLA-II ligands, since identification by MS has demonstrated highest fidelity for ligands with real MHC II binding (*41*). For HLA-I, only 4 peptides were detected in 2 tumors, all remaining were unique to one specific tumor. A small overlap existed with two published GBM data sets, suggesting that the ligandome of CNS tumors is more closely related than that of extracranial cancers. No viral, or retroviral sequences as suggested (*10*), could be identified in the AT/RT immunopeptidome.

Our epitope discovery platform additionally included the search for peptides derived from non-canonical protein translation. Such false or aberrant translation events result in small and extremely short-lived intracellular proteins, which are hard to detect by conventional proteomics but can nevertheless enter the proteasomal pathway. Intriguingly, half-life of a cryptic pMHC complex is about 90 times longer than that of the cytosolic source protein (*42*). Cryptic peptides have recently been reported to make up to 15% of the HLA class-I ligandome in normal as well as in malignant cells (*13, 14*). HLA-class-II cryptic peptides are unlikely to exist since class-II epitopes are longer and derived from large and stable internalized extracellular proteins (*43*). Cryptic peptides might represent a new and preferential source of targetable antigens for cancer immunotherapy (*31*), as they are abundant, tumor-specific and possibly shared between different tumors (*43*). Since they are most likely not part of the library of self-peptides presented during thymic T-cell selection, they can be regarded as non-mutated neoantigens. We were able to identify 61 tumor-specific, HLA-binding cryptic peptides not being present in the benign dataset (*17*), thus, more than half of the 116 HLA-class-I peptides in AT/RTs originated from non-canonical source proteins. Out-of-frame translations and gene products from 5’UTR regions dominated as reported previously (*14*). 33% of these peptides were repeatedly found either in several AT/RT samples or in a published GBM dataset (*13*). Notably, the peptide repertoire overlap between AT/RT and GBM was higher for cryptic than for natural HLA ligands. HLA-I binding prediction showed that ligandome peptides from natural proteins had significantly stronger HLA-binding than cryptic peptides, indicating that the former have undergone a more stringent intracellular selection process. Noteworthy, when we combined peptide ligands from all three subtypes suitable ligands were found for all patients with a median of 13 peptides per tumor (range 3-34), indicating that even low-mutational pediatric tumors give rise to actionable tumor-associated T-cell targets.

In contrast to other studies, which detected small sets of entity-spanning antigens in hematological malignancies (*40*) and glioblastomas (*44*), we could not identify network clusters for HLA-I or -II ligands using gene ontology tools. Since these algorithms were trained on larger data sets, it is possible that we missed them due to the rather low number of source proteins in our data (< 140). However, the allocation of source proteins to specific cellular compartments followed clear patterns. As expected, source proteins for natural class I ligands were mainly located in the cytoplasm, whereas class II peptides derived from membrane bound or intracellular vesicle associated proteins. Cryptic peptides were annotated to the nucleoplasm and to a lesser extent to the trans-Golgi network, suggesting that cryptic peptides are representatives of this tumor cell compartment on the cell surface. Thus, we postulate that cryptic peptides are an important component of the immunological homunculus of the AT/RT cell (Fig. 7). However, in contrast to its fixed neurological counterpart, the immunological homunculus is most likely highly dynamic, since inflammation (*45*), but also therapeutic elements such as chemotherapy or kinase inhibitors can profoundly impact on HLA expression and unmask new peptide epitopes (*46*). Since the cryptic ligandome of only few tumors has been unveiled so far, we do not know whether the rather high contribution of cryptic peptides to the ligandome is an AT/RT-specific phenomenon and related to the epigenetic dysregulation of this tumor, or whether frequencies of >50% can be found in other tumors as well. Certainly, the stability and composition of the immunopeptidome during therapy and disease progression requires more active investigation.

**Fig. 7.**
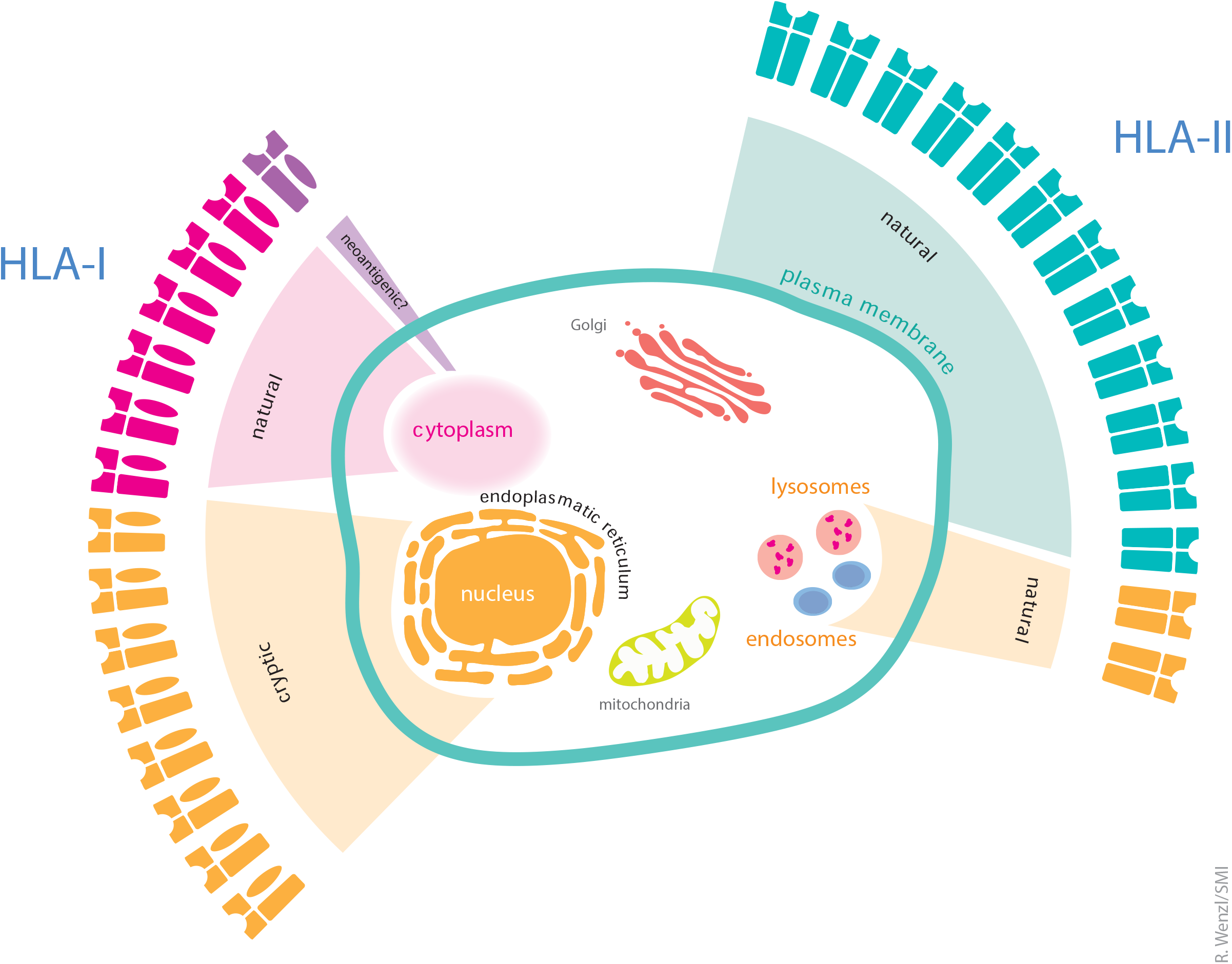
The immunological homunculus of AT/RTs. Illustration of the immunological representation of different cellular compartments of the AT/RT cell on HLA class-I (left) and -II molecules (right). Portion of the respective wings reflects the contribution of each peptide source to the class-I or -II peptidome. The size of the compartments in the outer circle corresponds to the representation of the compartment in the peptidome.

A basic question in tumor vaccine design is whether selected peptides can give rise to T-cell responses *in vivo*. We tried to address this issue on two levels: the level of *de novo* naïve T-cell priming (mimicking a vaccine response) and on the level of existing recall responses in AT/RT patients. For level one testing, we chose a validated T-cell assay in which highly purified naïve CD8+ T-cells were primed on peptide-loaded autologous dendritic cells (*19, 20*). In this assay, more than 80% of tested HLA-I peptides from natural, cryptic or neoantigenic origin yielded at least one positive response, suggesting that all three peptide sources are suitable for vaccine design. AT/RT patients showed no memory responses against the identified class-I peptides, indicating that patients are indeed ‘CD8 naïve’ for all three types of HLA-class I peptides investigated here. Non-canonical peptides were also not recognized in tumor-infiltrating CD8^+^ T cells in melanoma (*43*). It is conceivable that due to their exceptional short half-life cryptic peptides are not presented to T cells via dendritic cells in lymphatic tissue, i.e. that the peripheral T-cell repertoire of AT/RT patients is devoid of these specificities, but these could be *de novo* primed by therapeutic vaccines. In contrast to missing responses against class-I peptides, ~83% of the tested HLA-II epitopes were recognized by patients’ T_helper_ cells. This can partly be explained by the higher promiscuity of class-II ligands for HLA-II binding and presentation. Yet, this reactivity was tumor-specific since PBMCs of high-grade glioma patients recognized only ~50% of the tested HLA-II epitopes at a lower level whereas CD4-T-cells from healthy donors showed almost no reactivity. This indicates that the T_helper_ cell repertoire of AT/RT patients has been sensitized to tumor-associated class-II epitopes during disease initiation. Furthermore, our data show a partial overlap of immunopeptidomes of two CNS tumors, glioblastoma and AT/RTs on the level of natural and cryptic class-I epitopes as well as by shared CD4^+^ recall responses. 18% of the identified cryptic peptides were shared between AT/RTs and GBM, making these antigens highly promising candidates for future vaccines.

A limitation of our study certainly is that we worked predominantly with tumor banking material, which might have resulted in lower peptide yield due to suboptimal quality of material. Furthermore, the lack of clinical information to these samples precluded an analysis between immunological findings and clinical outcome.

In summary, AT/RTs as a paradigmatic example of a low-mutational pediatric cancer present a plethora of tumor-associated, immunogenic HLA-class-I and class-II peptides. More than half of the class-I peptides originated from non-canoncial translated proteins. Thus, personalized therapeutic vaccine cocktails should contain peptides from all three subgroups in order to have full representation of the tumor immunopeptidome in the vaccine. Future research has to clarify whether the cryptic class-I or natural class-II ligandome is enriched in epitopes, which span several CNS-tumor entities and could serve as a warehouse peptide pool.

## Methods

### Patients, tissues and blood samples

AT/RT tissue samples (n=18) were obtained from tumor banks at the University Hospitals in Würzburg, Bonn, Heidelberg, Berlin, and Tübingen. In n=5 patients treated in Tübingen and Würzburg, fresh tissue could be obtained directly from neurosurgical procedures. Informed consent from the guardians of the patients was obtained following internal regulations of the institutional review board (Ethics Commission at the Faculty of Medicine, University of Würzburg, #86/15). According to the requirements of the German HIT-Netzwerk, AT/RT diagnoses were centrally reviewed and confirmed in all 23 available cases by the DGNN reference neuropathology in Bonn. Tumour tissue samples have been snap frozen and stored in liquid nitrogen until usage. All samples were subject to ligandome analysis as described below. In n=10 cases, reference DNA from peripheral mononuclear cells was available, thus, in these cases whole exome sequencing and neoepitope identification could be performed. In 9 tumor samples, MPLA sequencing of the SMARCB1 locus was carried out to identify the nature of the SMARCB1 mutation. The whole workflow of AT/RT tissue workup is detailed in figure S1. For immunohistochemistry, n=17 additional formalin-fixed and paraffin-embedded (FFPE) AT/RT tumor tissues from the neuropathologies in Würzburg and München were used. Population coverage analysis for HLA class I and II was performed using the IEDB analysis resource (http://tools.iedb.org/population/)(*15*).

### HLA typing

HLA alleles for loci A, B, C, DRB1 and DQB1 were determined by standard sequencing procedures (sequence-based typing SBT, Protrans GmbH, Hockenheim, Germany or GenDx, The Netherlands) which provide high resolution typing. Confirmatory typing was undertaken by sequence-specific oligonucleotides (SSO, One Lambda or Olrup CareDx, San Francisco, USA) according to institutional regulations.

### Immunohistochemistry

Paraffin-embedded samples (thickness 2 µm) of n=17 AT/RT tissue were deparaffinized with Xylol and Ethanol and subsequently stained with standard immunohistochemistry procedures against CD3 (dilution 1:150), HLA class I (clone EMR 8-5, Abcam, dilution 1:1600), HLA class II (clone CR3/43, Abcam, dilution 1:1000), and PD-L1 (dilution 1:500). One day after primary antibody incubation, slides were stained with streptavidin-peroxidase-complex (BioGenex, Fremont CA, USA) for 10 min andd counterstained with Mayer’s hemalaun solution. Human tissue from tonsil (CD3, HLA class II), liver (HLA class I), and placenta (PD-L1) served as a positive control. Expression levels were evaluated semiquantitatively by two independent investigators and given as the frequency of positive cells per visual field. 10 visual fields per tumor were analyzed. For determining the PD-L1 expression level, the harmonized Cologne scoring system was used (*47*).

### Isolation of HLA ligands

HLA class I and clas II molecules were isolated using standard immunoaffinity purification (*48, 49*). In brief, snap-frozen AT/RT tissue samples were minced in lysis buffer containing 10 mM CHAPS/PBS (AppliChem, St. Louis, MO, USA/Gibco, Carlsbad, CA, USA) and one protease inhibitor tablet (Complete; Roche, Basel, Switzerland). We employed the pan-HLA class I-specific antibody W6/32 (*50*) and the HLA-DR-specific antibody L243 (*51*), both produced in house (University of Tübingen, Department of Immunology) from HB-95, and HB-55 hybridoma cell lines (ATCC, Manassas, VA) respectively. Furthermore, the pan-HLA class II-antibody Tü39 was employed (*52*), and produced in house from a hybridoma clone previously described. Antibodies were covalently linked to to CNBr-activated Sepharose (GE Healthcare, Chalfont St Giles, UK). HLA molecules were purified by cyclically passing the cleared protein lysate from AT/RT tissue over two columns. The first column contained 1 mg of W6/32 antibody coupled to sepharose, whereas the second column contained equal amounts of Tü39 (0.5 mg) and L243 (0.5 mg) antibody coupled to sepharose HLA-DR1 is known to be expressed at higher levels than the other class II allotypes, therefore the specific L243 mAb at the given stoichiometry was used (*53*). Tü39 is utilized complementarily to pull-down the remaining class II complexes. HLA–peptide complexes were eluted by repeated addition of 0.2% TFA (trifluoroacetic acid, Merck, Whitehouse Station, NJ, USA). Eluted HLA ligands were purified by ultrafiltration using centrifugal filter units (Amicon; Millipore, Billerica, MA, USA) with 3 kDa and 10 kDa pore size for HLA class I and HLA class II ligands, respectively. HLA ligands were desalted using ZipTip C18 pipette tips (Millipore). Extracted peptides were eluted in 35 µl 32.5% acetonitrile (Merck)/0.2% TFA, and vacuum-centrifuged to 2 – 5 µl and resuspended in 25 µl of 1% acetonitrile/0.05% TFA. Samples were stored at −80 °C until analysis by LC–MS/MS.

### Analysis of HLA ligands by LC-MS/MS

Peptide samples were separated by reversed-phase liquid chromatography (nano-UHPLC, UltiMate 3000 RSLCnano; Thermo Fisher, Waltham, MA, USA) and analyzed in online-coupled LTQ Orbitrap XL mass spectrometer (Thermo Fisher). Samples were analyzed in three or five technical replicates. Sample shares of 20% were trapped on a 75 µm × 2 cm trapping column (Acclaim PepMap RSLC; Thermo Fisher) at 4 µl/min for 5.75 min. Peptide separation was performed at 50 °C and a flow rate of 175 nl/min on a 50 µm × 25 cm separation column (Acclaim PepMap RSLC; Thermo Fisher) applying a gradient ranging from 2.4 to 32.0% of acetonitrile over the course of 90 min. Samples were analyzed on an LTQ Orbitrap XL using a top five CID (collision-induced dissociation) method with survey scans at 60k resolution and fragment ion detection in the ion trap (ITMS2) operated at normal scan speed. Fragmentation of HLA class I ligands was limited to precursors with a mass range of 400–650 m/z and charge states 2+ and 3+. For HLA class II ligands, a mass range of 300–1500 m/z with charge states ≥ 2 were selected for fragmentation.

### Database search

The software Proteome Discoverer (v.1.3; Thermo Fisher) was used to perform database search using the Mascot search engine (Mascot 2.2.04; Matrix Science, London, UK). The human proteome as comprised in the Swiss-Prot database (http://www.uniprot.org, status September 2013, 20,225 reviewed sequences contained) was used as reference database. The search combined data of technical replicates and was not restricted by enzymatic specificity. Precursor mass tolerance was set to 5 ppm, and fragment mass tolerance to 0.5 Da. Oxidation of methionine residues was allowed as a dynamic modification. False discovery rate (FDR) estmation was performed using the Percolator node (*54*) and was set to a peptide spectrum match level FDR ≤ 5%. For HLA class I ligands, peptide lengths were limited to 8–12 amino acids (AA), for HLA class II, peptides were limited to 8–25 AA of length. Protein grouping was disabled, allowing all proteins that contain a given peptide to be annotated as peptide source proteins Only HLA class I ligands mapping into one source protein alone were considered for this analysis (3,244 potential targets). As a final step of quality control, binding prediction was performed employing NetMHC-3.0 with a cut-off score of % Rank<0.5 for strong binders and <2 for weak binders (*55*).

### Whole genome DNA sequencing

Library preparation and whole genome sequencing of tumor and matching normal DNA from peripheral blood was carried out as described previously (*56*). Briefly, 1-5 µg of genomic DNA was fragmented to ~300 bp insert size using a Covaris device. Size selection was conducted by agarose gel excision. For Paired-end library preparation we used Illumina, Inc. v2 protocols. Sequencing was performed on HiSeq2000 machines at the German Cancer Research Center (DKFZ) Genomics and Proteomics Core Facility. Sequencing reads were mapped and aligned as describe (*3*). Briefly, we called variants using samtools and vcftools on the tumor bam files. The called positions were looked up in the control bam file and the resulting raw calls were filtered using different annotations and criteria to create a set of high confidence somatic SNVs. Indels were called using the pipeline platypus (*57*). All coordinates used in the analyses are based on human reference genome assembly hg19, GRCh37 (ncbi.nlm.nih.gov/assembly/ 2758). Gene annotations are based on genecode annotation release 19 (gencodegenes.org/releases/19.html). Whole Genome Sequencing data have been deposited at the European Genome-phenome Archive, EGA Study Accession IDEGAS00001001297.

The obtained mutated and the corresponding wildtype sequences were verified in Uniprot or Ensemble databases (https://www.uniprot.org/, http://apr2018.archive.ensembl.org/index.html). For each mutation a virtual 9mer-library was created moving the mutated amino acid from pos 1-9 and matched with the individual HLA class I type (HLA-ABC) on the NetMHCpan 4.0 server (http://www.cbs.dtu.dk/services/NetMHCpan). For each mutation the best binding peptide was selected according to binding affinity and %Rank.

### Immunogenicity prediction tools

For *in silico* immunogenicity prediction two different online platforms were used. VaxiJen2.0 is a publicly available online platform which uses an alignment-independent method for vaccine subunit prediction based on auto cross covariance transformation (http://www.ddg-pharmfac.net/vaxijen/VaxiJen/VaxiJen.html). Amino acid properties (hydrophobicity, size and polarity) are transcribed in z descriptors independently from the corresponding HLA allele (*58*). Protein sequences of source proteins were entered in fasta format, and *Tumour* selected as target organism. As suggested before, we chose a threshold of 0.45 to define probable protective antigens, slightly higher sensitivity than initially proposed threshold of 0.5(*23*). In order to consider also HLA-restriction, we additionally chose the IEDB immunogenicity prediction tool (http://tools.iedb.org/immunogenicity) (*59*). Since the original validation of this algorithm was performed only for nonamer-peptides were restricted our analysis to 9-mers, and to the HLA-molecules specified in the menu. Peptide sequences of potential epitopes were entered, and the T-cell recognition score was determined with a threshold of 0.1 was set to distinguish whether HLA-ligands were predicted to be “immunogenic” or “non-immunogenic.” Additionally, the mean T-cell recognition score was calculated for ligandome class I, ligandome cryptic, and neoantigenic peptides in order to compare immunogenicity of these three distinctive groups. Furthermore, the frequencies of probably protective/immunogenic antigens was calculated for the whole group as well as for the subgroups of strong, weak, or non-binders according NetMHC alogithms.

### Gene ontology and protein network analysis

Gene ontology and functional annotations of source proteins were defined and analyzed using DAVID Bioinformatics Resources 6.8, NIAID/NIH (https://david.ncifcrf.gov/summary.jsp) (*60*). Protein-protein interaction networks and functional enrichment analysis of proteins from different peptide sources was performed using STRING (https://string-db.org/cgi/input.pl)(*61*). For description of protein domains the PFAM 32.0 database was chosen (https://www.pfam.xfam.org). Overlap of ligandome source proteins was analyzed and visualized using the BioVenn web application (https://www.biovenn.nl) (*62*).

### Naïve CD8^+^ T-cell priming and T-cell expansion protocol

For T-cell priming assays we used a previously described protocol (*19, 20*). In brief, naïve CD8^+^ T cells were isolated from the peripheral blood of healthy donors positive for HLA-A02*01 (n=4) or -B07*02 (n=1) using a purification of CD8^+^ events, followed by depletion with CD45RO and CD57 microbeads on LD and LS columns (Miltenyi, Bergisch Gladbach, Germany). Isolated naïve CD8^+^ T cells were incubated with IL-7 overnight. The next day, T cells were washed and mixed with irradiated, matured autologous DCs, pulsed with either the heteroclitic HLA-A02*01-restricted peptide Melan-A26-35L as a control or the investigational peptides as indicated at 1 µg/ml. IL-21 (30 ng/ml, Peprotech) was added to the co-culture on day 0 the next morning. IL-7 and IL-15 (5 ng/ml, Peprotech) were added on days 3, 5, and 7 and cells were harvested on day 10, some of them after restimulation with the original peptide. The Melan-A26-35L (ELAGIGILTV), and investigational peptides were purchased from jpt (Berlin, Germany). Harvested T-cells on day 10 or 12 were analyzed for TNFα/IFNγ-positivity. T-cell responses three times above the negative control (autologous DCs without peptide) were considered positive, those above the background but <3x were considered intermediate.

T-cell responses defined by specific IFNγ-secretion were measured using a stimulation-expansion-restimulation protocol (*63*): PBMCs were isolated by Ficoll, stimulated with mature, peptide-loaded autologous DCs at a 4:1 (T:DC) ratio and expanded subsequently with IL-15 (5 ng/ml) for 12–14 days. Medium and IL-15 was refreshed every second or third day. On days 7 and 14 PBMCs were restimulated with the same peptide loaded DCs. Negative control was stimulation with empty DCs (mature, autologous DCs without peptide loading). 6 h after the last restimulation, T cells were analyzed for IFNγ-production by intracellular cytokine staining. Only T-cell responses three times above the negative control (autologous DCs without peptide) were considered positive. In HLA class I assays, the CEF-pool peptide mix (jpt, Hamburg, Germany) was used as a positive control.

### Identification of cryptic peptides

Cryptic HLA-I peptides were identified using Peptide-PRISM as described recently (*13*). De novo peptide sequencing was performed with PEAKS X (Bioinformatics Solutions Inc., Canada) (*64*). Raw data refinement was performed with the following settings: (i) Merge Options: no merge; (ii) Precursor Options: corrected; (iii) Charge Options: no correction; (iv) Filter Options: no filter; (v) Process: true; (vi) Default: true; (vii) Associate Chimera: yes. De novo sequencing was performed with Parent Mass Error Tolerance set to 10 ppm. Fragment Mass Error Tolerance was set to 0.15 Da, and Enzyme was set to none. The following variable modifications have been used: Oxidation (M), pyro-Glu from Q (N-term Q), and carbamidomethylation (C). A maximum of 3 variable post-translational modifications (PTMs) were allowed per peptide. Up to 10 de novo sequencing candidates were reported for each identified fragment ion mass spectrum, with their corresponding average local confidence (ALC) score. Because we applied the chimeric spectra option of PEAKS X, two or more TOP10 candidate lists could be assigned to a single fragment ion spectrum. Two tables (“all de novo candidates” and “de novo peptides”) were exported from PEAKS for further analysis.

All de novo sequence candidates were matched against the 6-frame translated human genome (hg38) and the 3-frame translated human transcriptome (ENSEMBL 90) using Peptide-PRISM. Results were filtered to 10% FDR for each category (CDS, UTR5, OffFrame, ncRNA, UTR3, intronic, intergenic). NetMHCpan 4.0 was used to predict binding affinities for all identified HLA-I peptides for all HLA alleles of the corresponding patient. The allele with the minimal rank reported by NetMHCpan was annotated. As per default, we used a cutoff of 0.5% rank for strong binders and 2% rank for weak binders.

Identification of cryptic peptides from the glioblastoma (GBM) data set (*18*) has been performed in the same way using Peptide-PRISM. Comprehensive results have been published recently (*13*).

Synthetic peptides were analyzed on an LTQ-Orbitrap Velos Pro (Thermo Scientific) equipped with a PicoView Ion Source (New Objective) and coupled to an EASY-nLC 1000 (Thermo Scientific). Peptides were loaded on capillary columns (PicoFrit, 30 cm x 150 µm ID, New Objective) self-packed with ReproSil-Pur 120 C18-AQ, 1.9 µm (Dr. Maisch) and separated with a 30-minute linear gradient from 3% to 30% acetonitrile and 0.1% formic acid and a flow rate of 500 nl/min. MS scans were acquired in the Orbitrap analyzer with a resolution of 30,000 at m/z 400, MS/MS scans were acquired in the Ion Trap analyzer with normal scan rate using CID fragmentation with 35% normalized collision energy. A TOP10 data-dependent MS/MS method was used; dynamic exclusion was applied with a repeat count of 1 and an exclusion duration of 120 seconds; singly charged precursors were excluded from selection. Minimum signal threshold for precursor selection was set to 20,000. AGC was used with AGC target with a value of 1e6 for MS scans and 1e4 for MS/MS scans. Lock mass option was applied for internal calibration in all runs using background ions from protonated decamethylcyclopentasiloxane (m/z 371.10124).

### Statistical analyis

For defining correlations between different IHC markers, the percentages of marker positive cells in neuropathological slides were logit-transformed and plotted by Pearson linear regression using SPSS V26. Differences between two groups were assessed by nonparametric Mann–Whitney U test assuming unequal variances. For comparison of multiple groups, the non-parametric Kruskal–Wallis rang-sum test was used. For differences within a group the two-tailed t test was applied. p < 0.05 was considered statistically significant.

## Data Availability

All online tools used in this study are publicly available online as indicated including the benign tissue database. Peptide sequence and source protein information are provided in the supplementary materials. The mass spectrometry proteomics data have been deposited to the ProteomeXchange Consortium (http://proteomecentral.proteomexchange.org) via the PRIDE partner repository (doi: 10.1093/nar/gky1106).

## Aknowledgement

We thank Robert Wenzl for graphic design support with Figure 7, Götz Gelbrich for aid with the statistical workup of IHC-data, and Stefan Stevanović for support with MS data analysis and critical comments on the project progress.

## Funding resources

EU-TRANSCAN JCT 2013 (ATRTPepVac), funding agency for Germany: Bundesministerium für Bildung und Forschung (BMBF), for Israel: Ministry of Health (MoH)

Parents Initiative Group for Children with Leukemia and Solid Tumors Würzburg e.V. Tour of Hope Foundation

Elfrieda-Albert-Stiftung

Vogel-Stiftung Dr. Eckernkamp

## Author contribution

Conceptualization: ME, HGR, YR

Methodology: AM, HGR, MW, ME

Investigation: AM, AS, NT, AK, PJ, JL, CM, LH, EK, MW, FO, AL

Provision of tumor material: JK, MS, UT, MEb, MF, TP

Visualization: AM, ME

Writing – original draft: AM, AS, ME

Writing – review & editing: ME, PGS, HGR, YR

## References

1. M. C. Fruhwald et al., Age and DNA methylation subgroup as potential independent risk factors for treatment stratification in children with atypical teratoid/rhabdoid tumors. Neuro-Oncology 22, 1006–1017 (2020).

2. I. Versteege et al., Truncating mutations of hSNF5/INI1 in aggressive paediatric cancer. Nature 394, 203–206 (1998).

3. P. D. Johann et al., Atypical Teratoid/Rhabdoid Tumors Are Comprised of Three Epigenetic Subgroups with Distinct Enhancer Landscapes. Cancer Cell 29, 379–393 (2016).

4. J. Torchia et al., Molecular subgroups of atypical teratoid rhabdoid tumours in children: an integrated genomic and clinicopathological analysis. Lancet Oncol 16, 569–582 (2015).

5. B. Vogelstein et al., Cancer genome landscapes. Science 339, 1546–1558 (2013).

6. J. D. Fumet, C. Truntzer, M. Yarchoan, F. Ghiringhelli, Tumour mutational burden as a biomarker for immunotherapy: Current data and emerging concepts. Eur J Cancer 131, 40–50 (2020).

7. J. Q. Lu, B. A. Wilson, V. W. Yong, J. Pugh, V. Mehta, Immune cell infiltrates in atypical teratoid/rhabdoid tumors. Can J Neurol Sci 39, 605–612 (2012).

8. V. Melcher et al., Macrophage-tumor cell interaction promotes ATRT progression and chemoresistance. Acta Neuropathol 139, 913–936 (2020).

9. H. E. Chun et al., Identification and Analyses of Extra-Cranial and Cranial Rhabdoid Tumor Molecular Subgroups Reveal Tumors with Cytotoxic T Cell Infiltration. Cell Rep 29, 2338–2354 e2337 (2019).

10. A. Leruste et al., Clonally Expanded T Cells Reveal Immunogenicity of Rhabdoid Tumors. Cancer Cell 36, 597–612 e598 (2019).

11. Y. Grabovska et al., Pediatric pan-central nervous system tumor analysis of immune-cell infiltration identifies correlates of antitumor immunity. Nature communications 11, 4324 (2020).

12. S. W. van Gool et al., Immunotherapy in atypical teratoid-rhabdoid tumors: Data from a survey of the HGG-Immuno Group. Cytotherapy, (2016).

13. F. Erhard, L. Dolken, B. Schilling, A. Schlosser, Identification of the Cryptic HLA-I Immunopeptidome. Cancer Immunol Res, (2020).

14. C. M. Laumont et al., Global proteogenomic analysis of human MHC class I-associated peptides derived from non-canonical reading frames. Nature communications 7, 10238 (2016).

15. H. H. Bui et al., Predicting population coverage of T-cell epitope-based diagnostics and vaccines. BMC Bioinformatics 7, 153 (2006).

16. R. Vita et al., The immune epitope database (IEDB) 3.0. Nucleic Acids Research 43, D405–D412 (2015).

17. A. Marcu et al., The HLA Ligand Atlas - A resource of natural HLA ligands presented on benign tissues. bioRxiv, (2020).

18. B. Shraibman et al., Identification of Tumor Antigens Among the HLA Peptidomes of Glioblastoma Tumors and Plasma. Mol Cell Proteomics 17, 2132–2145 (2018).

19. M. Wolfl, P. D. Greenberg, Antigen-specific activation and cytokine-facilitated expansion of naive, human CD8+ T cells. Nat Protoc 9, 950–966 (2014).

20. M. Wolfl et al., Primed tumor-reactive multifunctional CD62L+ human CD8+ T cells for immunotherapy. Cancer Immunol Immunother 60, 173–186 (2011).

21. V. Dutoit et al., Exploiting the glioblastoma peptidome to discover novel tumour-associated antigens for immunotherapy. Brain 135, 1042–1054 (2012).

22. J. K. Peper et al., HLA ligandomics identifies histone deacetylase 1 as target for ovarian cancer immunotherapy. Oncoimmunology 5, e1065369 (2016).

23. M. G. Klatt et al., Carcinogenesis of renal cell carcinoma reflected in HLA ligands: A novel approach for synergistic peptide vaccination design. Oncoimmunology 5, (2016).

24. B. Engels et al., Relapse or eradication of cancer is predicted by peptide-major histocompatibility complex affinity. Cancer Cell 23, 516–526 (2013).

25. M. Aleksic et al., Different affinity windows for virus and cancer-specific T-cell receptors: implications for therapeutic strategies. Eur J Immunol 42, 3174–3179 (2012).

26. P. A. Ott et al., An immunogenic personal neoantigen vaccine for patients with melanoma. Nature 547, 217–221 (2017).

27. M. Bassani-Sternberg et al., Direct identification of clinically relevant neoepitopes presented on native human melanoma tissue by mass spectrometry. Nature communications 7, 13404 (2016).

28. A. Newey et al., Immunopeptidomics of colorectal cancer organoids reveals a sparse HLA class I neoantigen landscape and no increase in neoantigens with interferon or MEK-inhibitor treatment. Journal for Immunotherapy of Cancer 7, (2019).

29. M. A. Purbhoo, D. J. Irvine, J. B. Huppa, M. M. Davis, T cell killing does not require the formation of a stable mature immunological synapse. Nature Immunology 5, 524–530 (2004).

30. K. A. Marijt, E. M. Doorduijn, T. van Hall, TEIPP antigens for T-cell based immunotherapy of immune-edited HLA class I-low cancers. Molecular Immunology 113, 43–49 (2019).

31. C. M. Laumont et al., Noncoding regions are the main source of targetable tumorspecific antigens. Sci Transl Med 10, (2018).

32. Q. C. Zhao et al., Proteogenomics Uncovers a Vast Repertoire of Shared Tumor-Specific Antigens in Ovarian Cancer. Cancer Immunology Research 8, 544–555 (2020).

33. R. S. Lee et al., A remarkably simple genome underlies highly malignant pediatric rhabdoid cancers. J Clin Invest 122, 2983–2988 (2012).

34. V. Vlkova et al., Epigenetic regulations in the IFN gamma signalling pathway: IFN gamma-mediated MHC class I upregulation on tumour cells is associated with DNA demethylation of antigen-presenting machinery genes. Oncotarget 5, 6923–6935 (2014).

35. F. Garrido, N. Aptsiauri, E. M. Doorduijn, A. M. G. Lora, T. van Hall, The urgent need to recover MHC class I in cancers for effective immunotherapy. Current Opinion in Immunology 39, 44–51 (2016).

36. M. W. Loffler et al., Mapping the HLA Ligandome of Colorectal Cancer Reveals an Imprint of Malignant Cell Transformation. Cancer Research 78, 4627-+ (2018).

37. A. Neumann et al., Identification of HLA ligands and T-cell epitopes for immunotherapy of lung cancer. Cancer Immunol Immunother 62, 1485–1497 (2013).

38. J. S. Stickel et al., Quantification of HLA class I molecules on renal cell carcinoma using Edman degradation. BMC Urol 11, 1 (2011).

39. T. Bilich et al., Mass spectrometry-based identification of a B-cell maturation antigen-derived T-cell epitope for antigen-specific immunotherapy of multiple myeloma. Blood Cancer J 10, (2020).

40. L. Backert et al., A meta-analysis of HLA peptidome composition in different hematological entities: entity-specific dividing lines and “panleukemia” antigens. Oncotarget 8, 43915–43924 (2017).

41. B. B. Chen et al., Predicting HLA class II antigen presentation through integrated deep learning. Nature Biotechnology 37, 1332-+ (2019).

42. M. V. Ruiz Cuevas et al., Most non-canonical proteins uniquely populate the proteome or immunopeptidome. Cell Rep 34, (2021).

43. C. E. Chong et al., Integrated proteogenomic deep sequencing and analytics accurately identify non-canonical peptides in tumor immunopeptidomes. Nature communications 11, (2020).

44. M. C. Neidert et al., The natural HLA ligandome of glioblastoma stem-like cells: antigen discovery for T cell-based immunotherapy. Acta Neuropathol 135, 923–938 (2018).

45. S. Murata, Y. Takahama, M. Kasahara, K. Tanaka, The immunoproteasome and thymoproteasome: functions, evolution and human disease. Nat Immunol 19, 923–931 (2018).

46. C. Y. Oh et al., ALK and RET Inhibitors Promote HLA Class I Antigen Presentation and Unmask New Antigens within the Tumor Immunopeptidome. Cancer Immunology Research 7, 1984–1997 (2019).

47. A. H. Scheel et al., Harmonized PD-L1 immunohistochemistry for pulmonary squamous-cell and adenocarcinomas. Mod Pathol 29, 1165–1172 (2016).

48. K. Falk, O. Rotzschke, S. Stevanovic, G. Jung, H. G. Rammensee, Allele-specific motifs revealed by sequencing of self-peptides eluted from MHC molecules. Nature 351, 290–296 (1991).

49. D. J. Kowalewski et al., HLA ligandome analysis identifies the underlying specificities of spontaneous antileukemia immune responses in chronic lymphocytic leukemia (CLL). Proc Natl Acad Sci U S A 112, E166–175 (2015).

50. C. J. Barnstable et al., Production of Monoclonal Antibodies to Group-a Erythrocytes, Hla and Other Human Cell-Surface Antigens - New Tools for Genetic-Analysis. Cell 14, 9–20 (1978).

51. J. M. Goldman, J. Hibbin, L. Kearney, K. Orchard, K. H. Thng, Hla-Dr Monoclonal-Antibodies Inhibit the Proliferation of Normal and Chronic Granulocytic-Leukemia Myeloid Progenitor Cells. Brit J Haematol 52, 411–420 (1982).

52. G. Pawelec, W. Newman, U. Schwulera, P. Wernet, Heterogeneity of Human Natural-Killer Recognition Demonstrated by Cloned Effector-Cells and Differential Blocking of Cyto-Toxicity with Monoclonal-Antibodies. Cellular Immunology 92, 31–40 (1985).

53. S. Boegel et al., HLA and proteasome expression body map. Bmc Med Genomics 11, (2018).

54. L. Kall, J. D. Canterbury, J. Weston, W. S. Noble, M. J. MacCoss, Semi-supervised learning for peptide identification from shotgun proteomics datasets. Nature methods 4, 923–925 (2007).

55. C. Lundegaard et al., NetMHC-3.0: accurate web accessible predictions of human, mouse and monkey MHC class I affinities for peptides of length 8-11. Nucleic Acids Research 36, W509–W512 (2008).

56. D. T. Jones et al., Recurrent somatic alterations of FGFR1 and NTRK2 in pilocytic astrocytoma. Nat Genet 45, 927–932 (2013).

57. A. Rimmer et al., Integrating mapping-, assembly-and haplotype-based approaches for calling variants in clinical sequencing applications. Nat Genet 46, 912–918 (2014).

58. I. A. Doytchinova, D. R. Flower, VaxiJen: a server for prediction of protective antigens, tumour antigens and subunit vaccines. BMC Bioinformatics 8, 4 (2007).

59. J. J. A. Calis et al., Properties of MHC Class I Presented Peptides That Enhance Immunogenicity. Plos Comput Biol 9, (2013).

60. W. Huang da, B. T. Sherman, R. A. Lempicki, Bioinformatics enrichment tools: paths toward the comprehensive functional analysis of large gene lists. Nucleic Acids Res 37, 1–13 (2009).

61. D. Szklarczyk et al., STRING v11: protein-protein association networks with increased coverage, supporting functional discovery in genome-wide experimental datasets. Nucleic Acids Res 47, D607–D613 (2019).

62. T. Hulsen, J. de Vlieg, W. Alkema, BioVenn - a web application for the comparison and visualization of biological lists using area-proportional Venn diagrams. BMC Genomics 9, 488 (2008).

63. U. S. Kammula et al., Functional analysis of antigen-specific T lymphocytes by serial measurement of gene expression in peripheral blood mononuclear cells and tumor specimens. J Immunol 163, 6867–6875 (1999).

64. J. Zhang et al., PEAKS DB: De Novo Sequencing Assisted Database Search for Sensitive and Accurate Peptide Identification. Molecular & Cellular Proteomics 11, (2012).

