## supplementary figure 1 for "Natural and cryptic peptides dominate the immunopeptidome of atypical teratoid rhabdoid tumors"

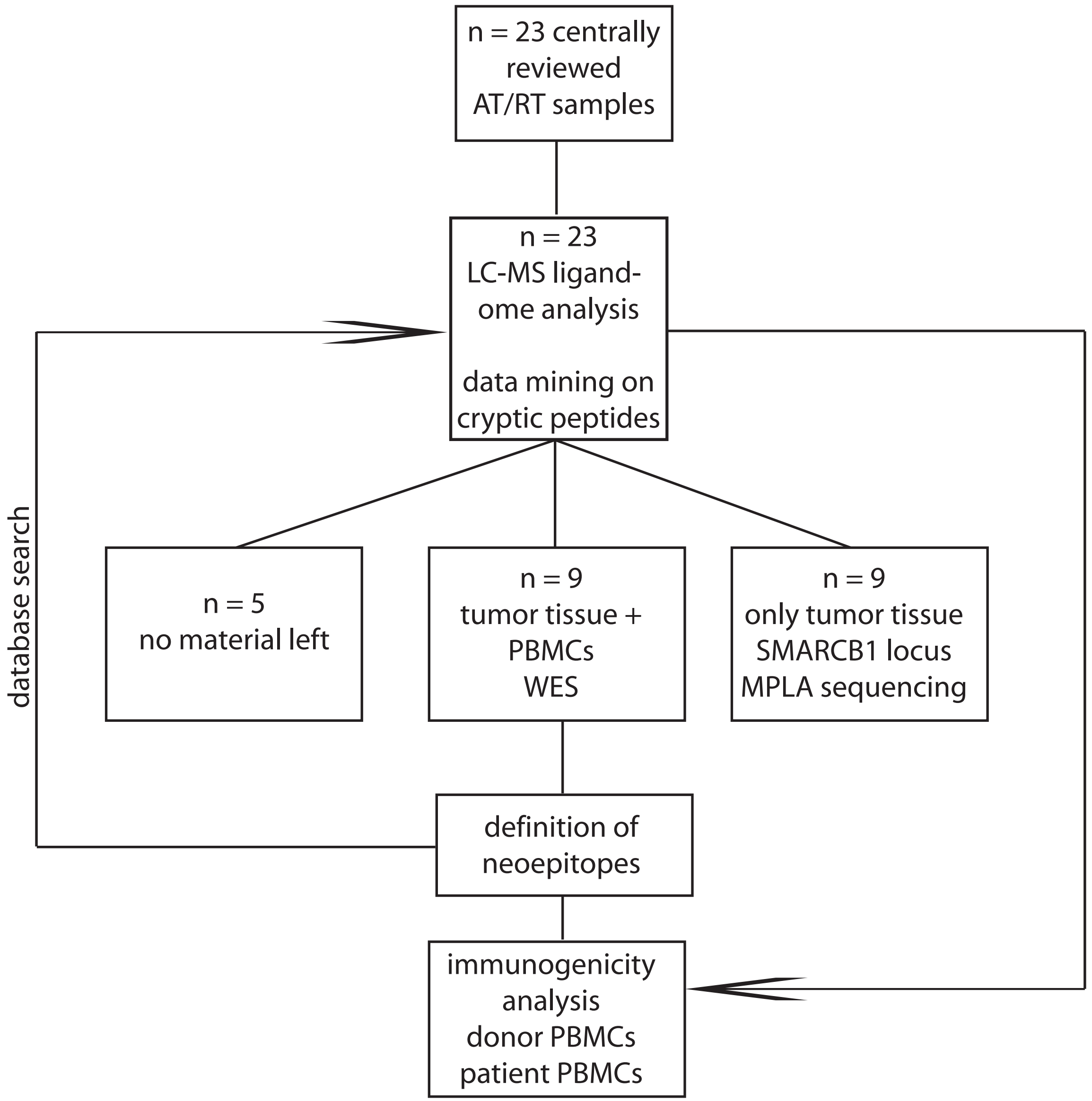


**Supplementary Figure 1:** Workflow of tumor tissue and PBMC samples from 23 patients with confirmed AT/RT.
