## supplementary figure 2 for "Natural and cryptic peptides dominate the immunopeptidome of atypical teratoid rhabdoid tumors"

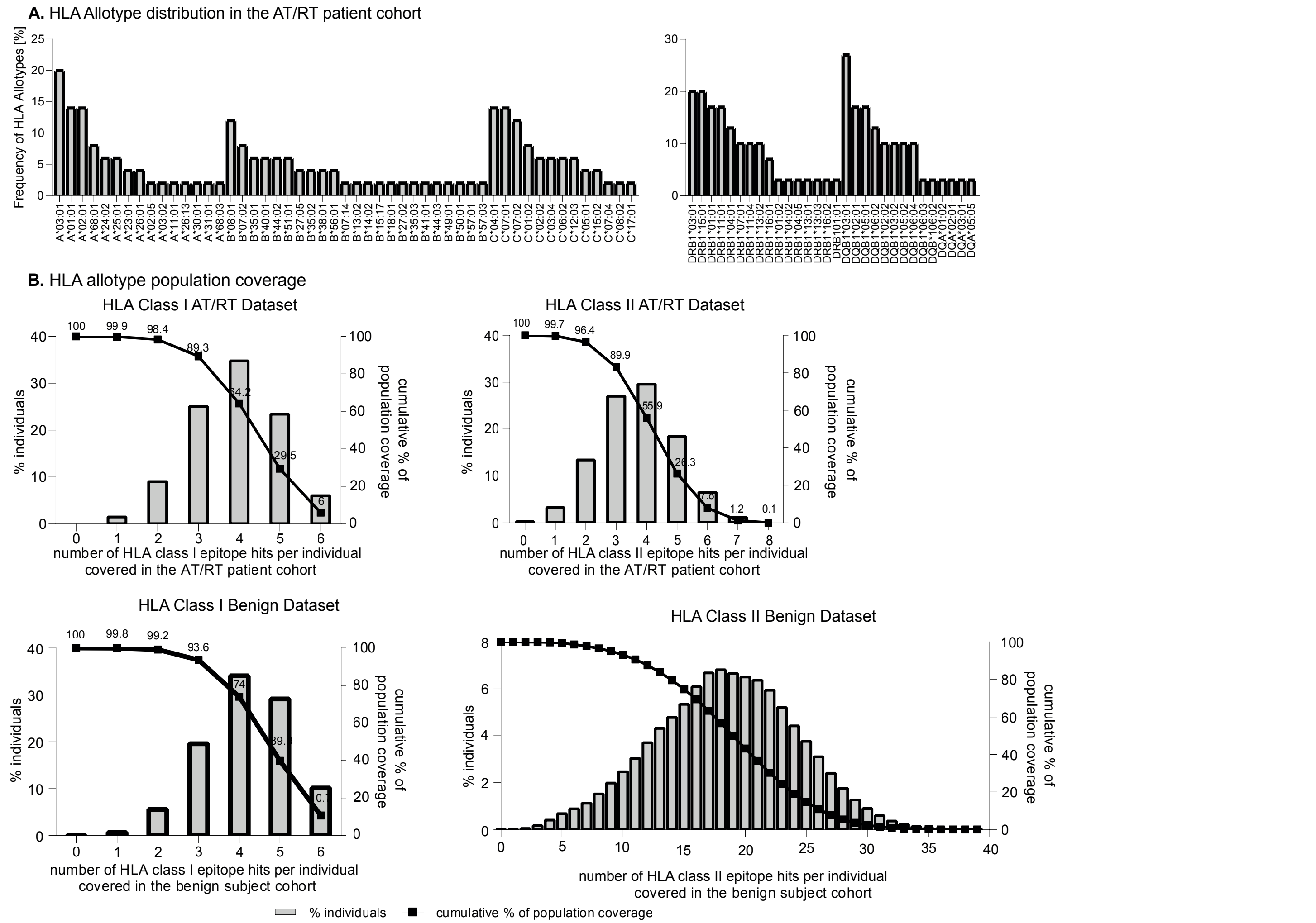


**Supplementary Figure 2:**
Distribution of the 51 HLA-A, -B, -C, DR, and 30 DQ allotypes within the AT/RT cohort (A). Population overage of HLA-allotypes was calculated for the AT/RT cohort (upper panels) and the comparative benign tissue dataset (lower panels) using the IEDB database (http://tools/iedb.org/population) (B). The cumulative coverage is depicted as a black line, whereas the percentage of indiviudals with a given number of epitope hits is shown as a column.
