## supplementary figure 3 for "Natural and cryptic peptides dominate the immunopeptidome of atypical teratoid rhabdoid tumors"

#### Supplementary Figure 3A

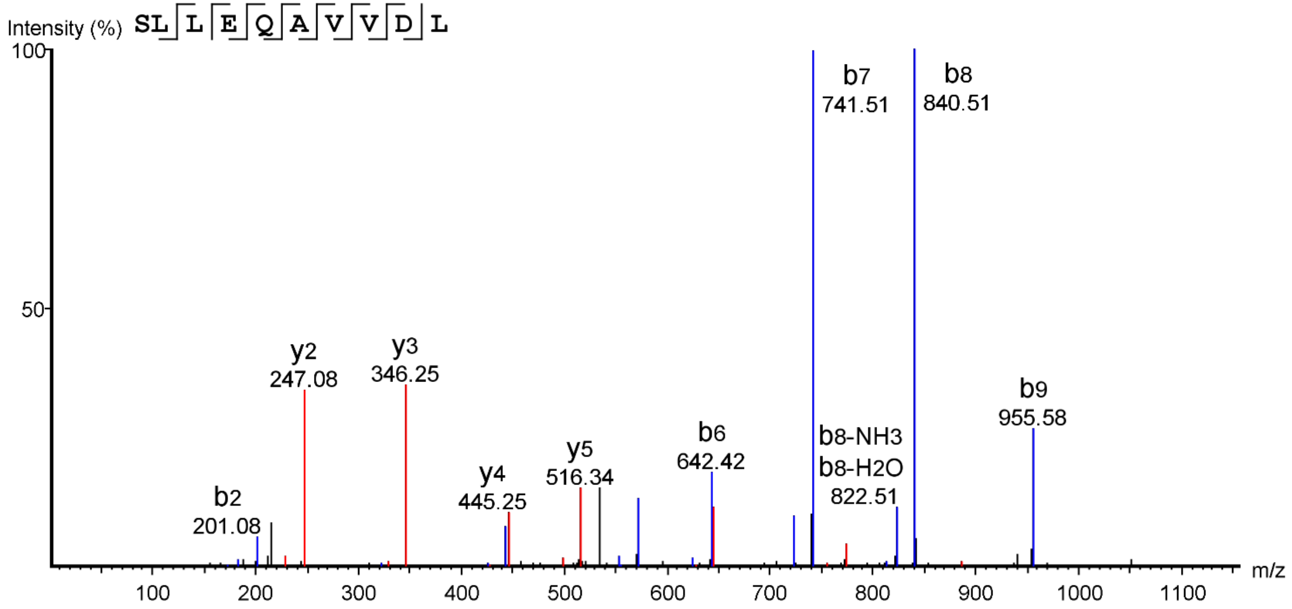


Synthetic reference peptide

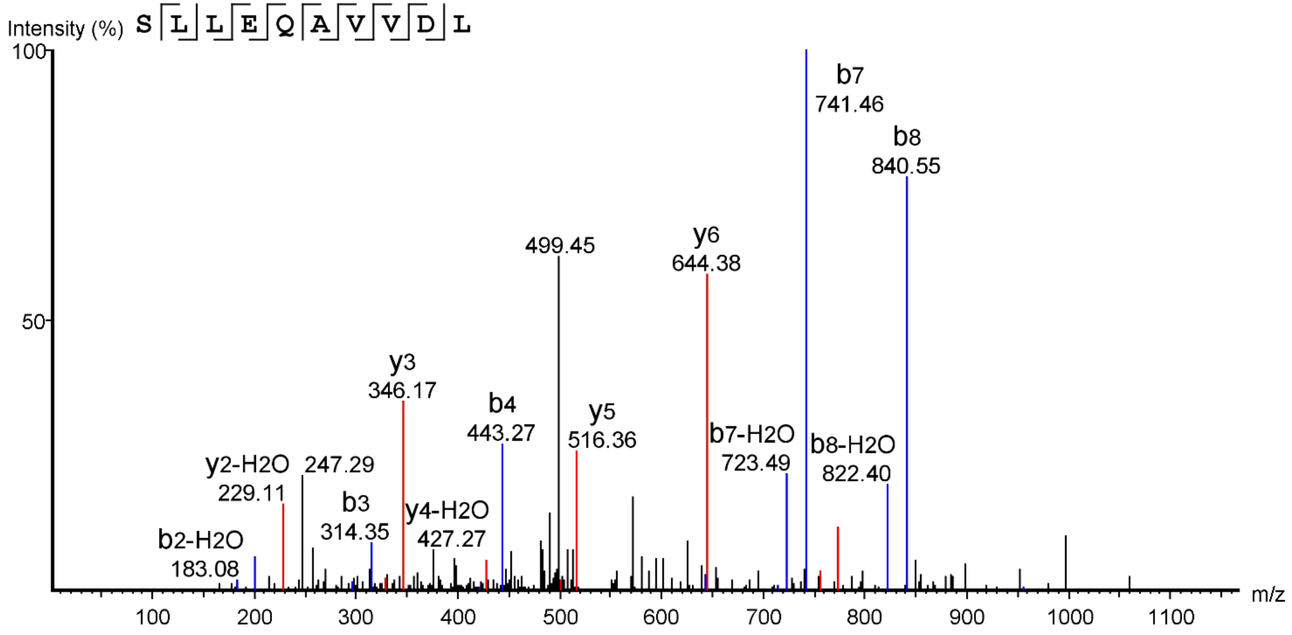


Original peptide

**Supplementary Figure 3B**

Synthetic reference peptide

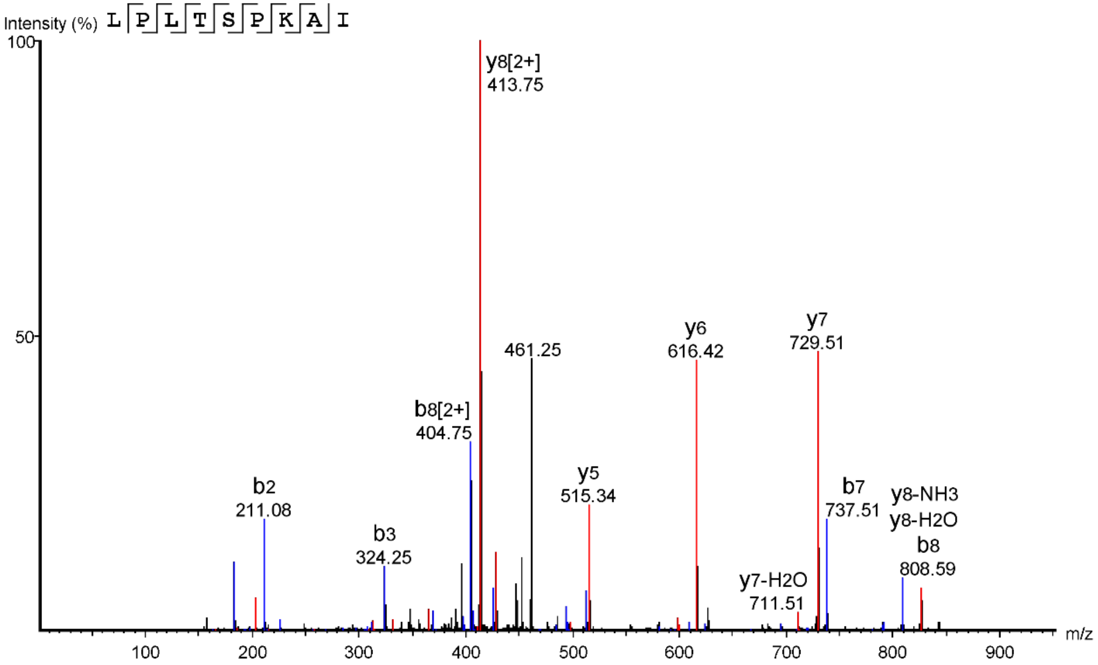


Original peptide

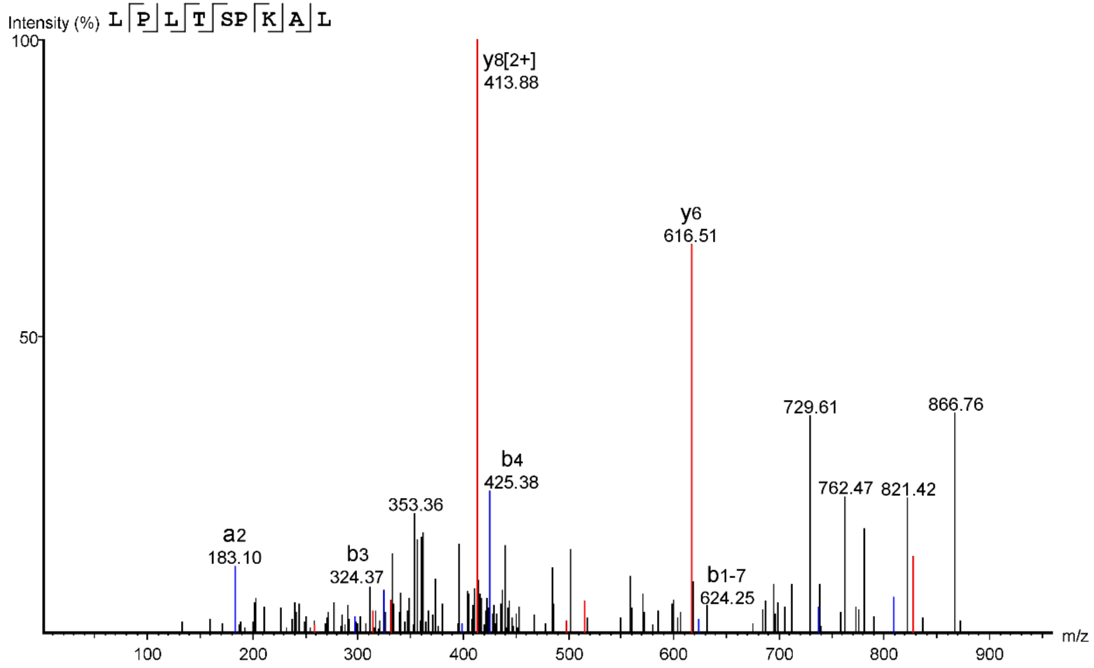

**Supplementary Figure 3C**

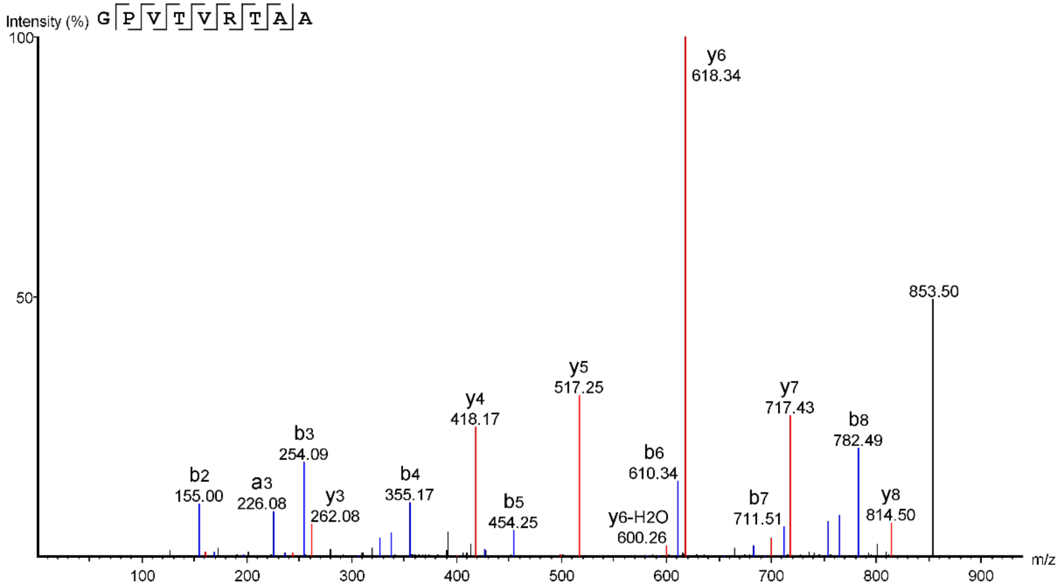


Synthetic reference peptide

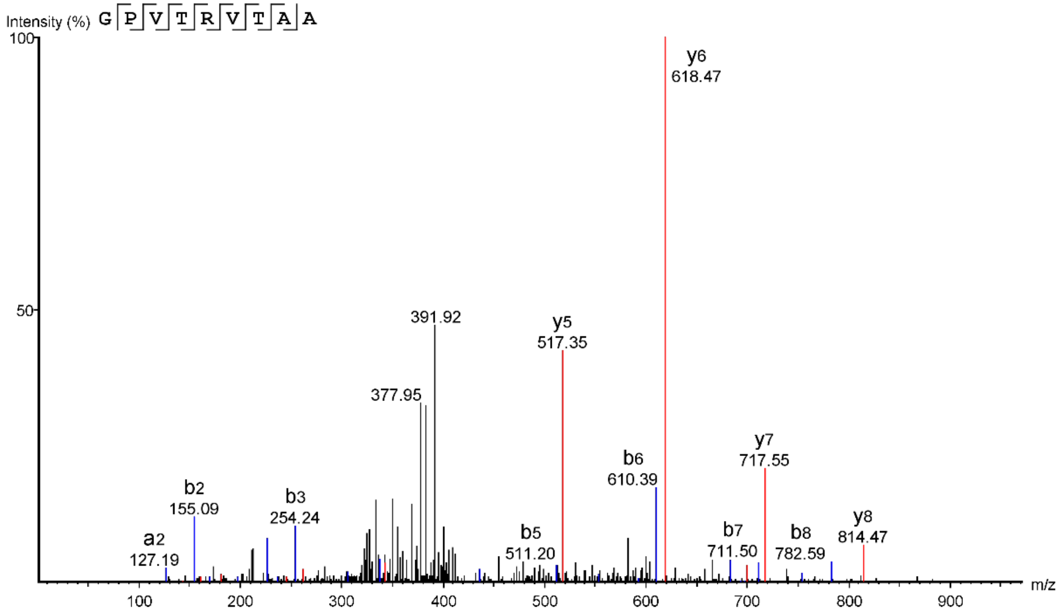


Original peptide

### Supplementary Figure 3D

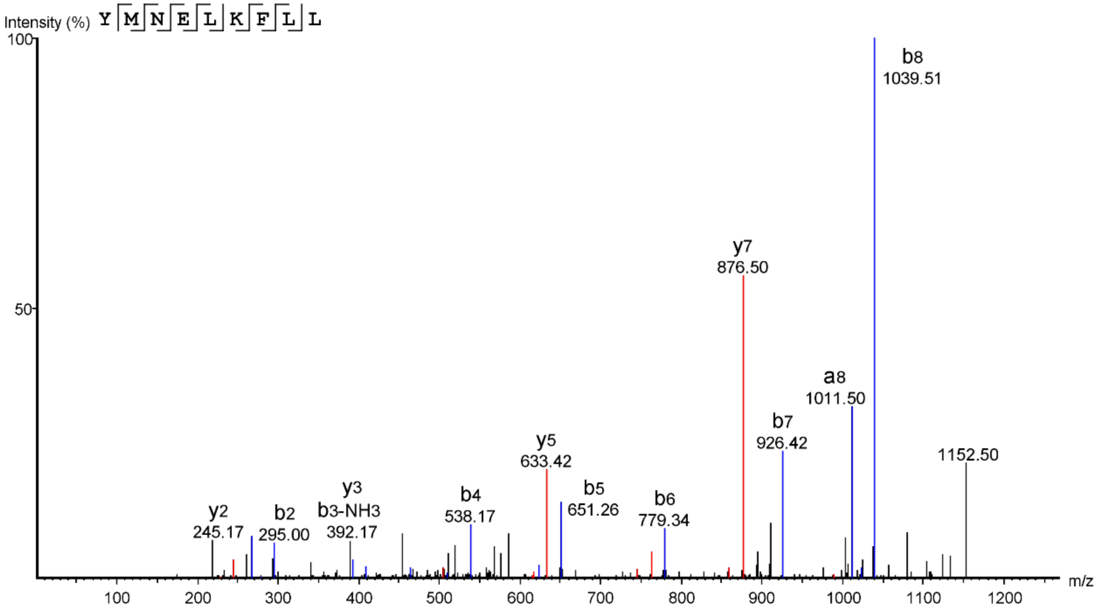


Synthetic reference peptide

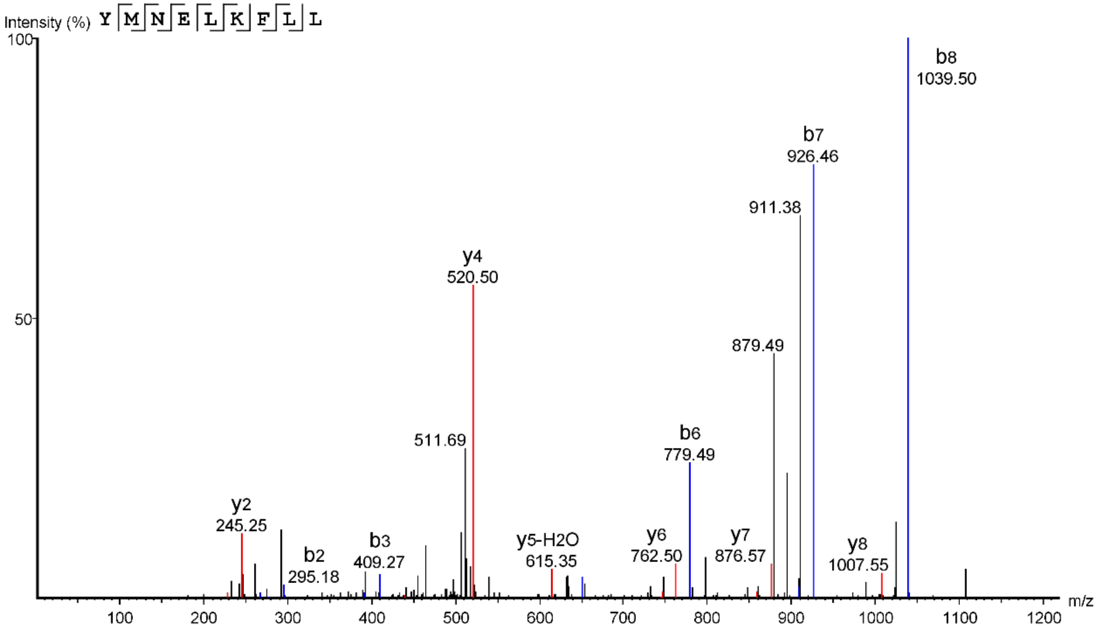


Original peptide

**Supplementary Figure 3E**

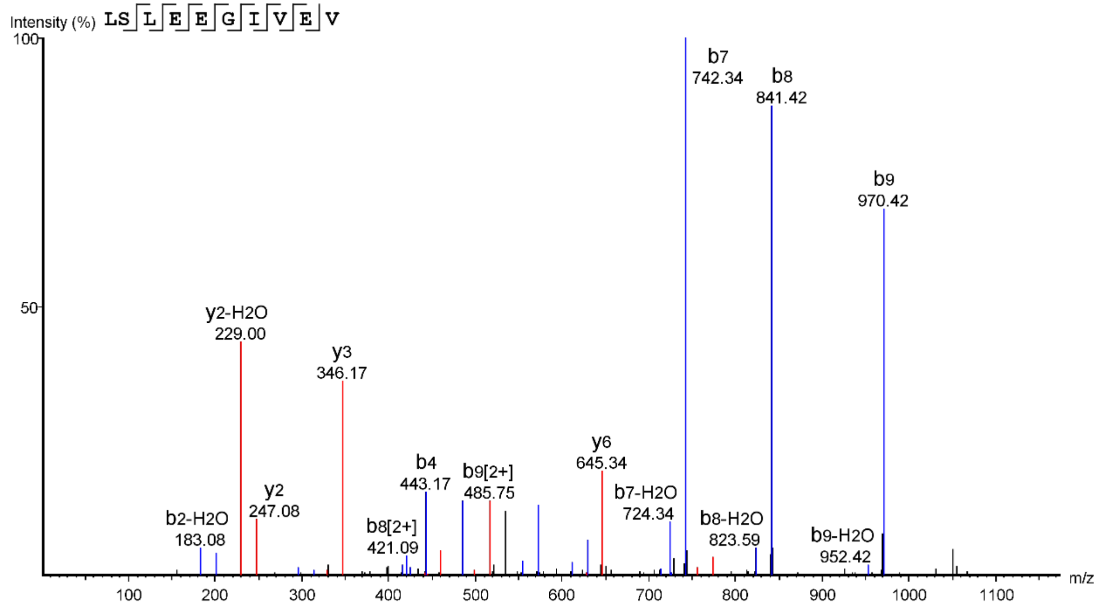


Synthetic reference peptide

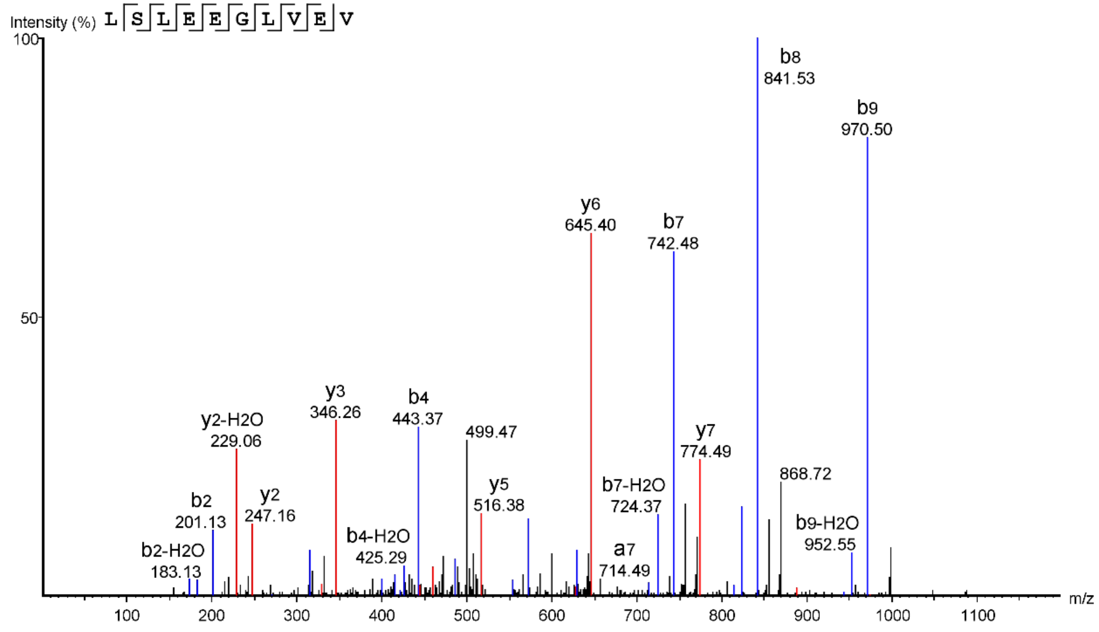


Original peptide

**Supplementary Figure 3F**

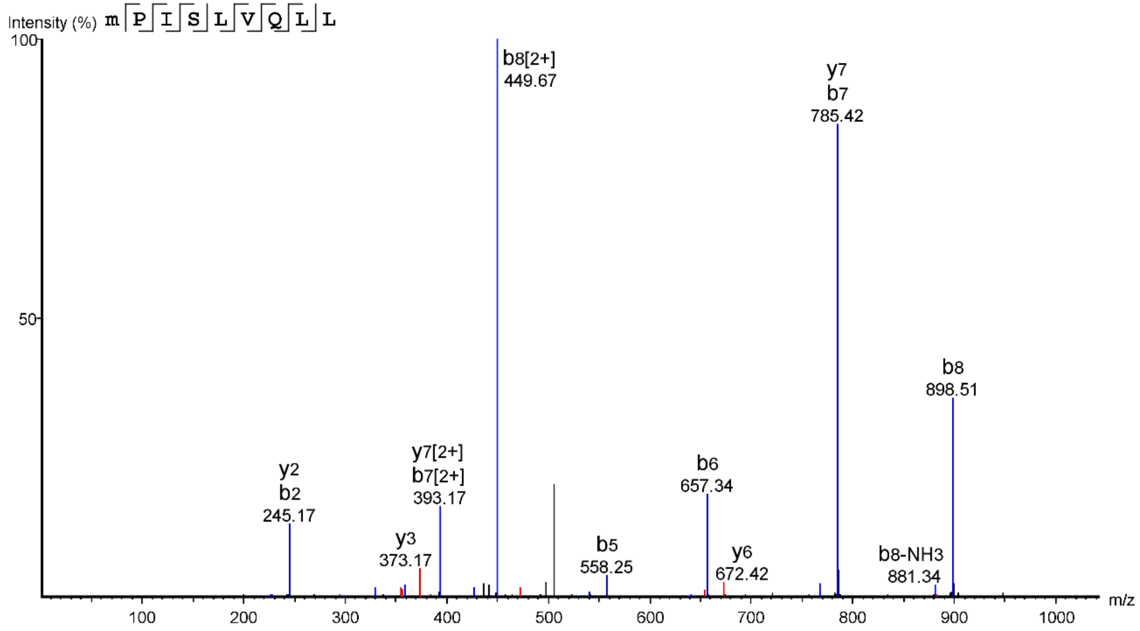


Synthetic reference peptide

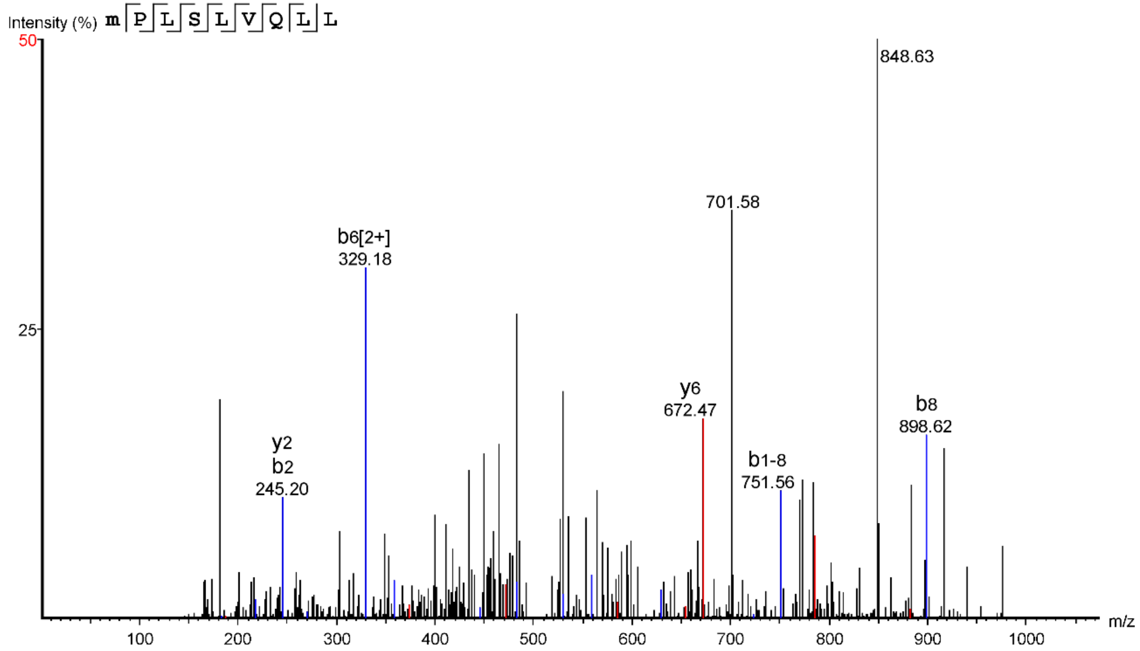


Original peptide

### **Supplementary Figure 3**

Supplementary Figure 3A-F: Fragment ion spectra of cryptic HLA-I peptides identified from AT/RT samples in comparison to reference spectra generated from the corresponding synthetic peptides. HLA peptides from AT/RT samples were analyzed on an Orbitrap XL applying collisional-induced dissociation (CID) in the ion trap. Reference spectra were analyzed on an Orbitrap Velos Pro applying ion trap CID. On both instruments fragment ion spectra were recorded with the ion trap.
