## supplementary figure 4 for "Natural and cryptic peptides dominate the immunopeptidome of atypical teratoid rhabdoid tumors"

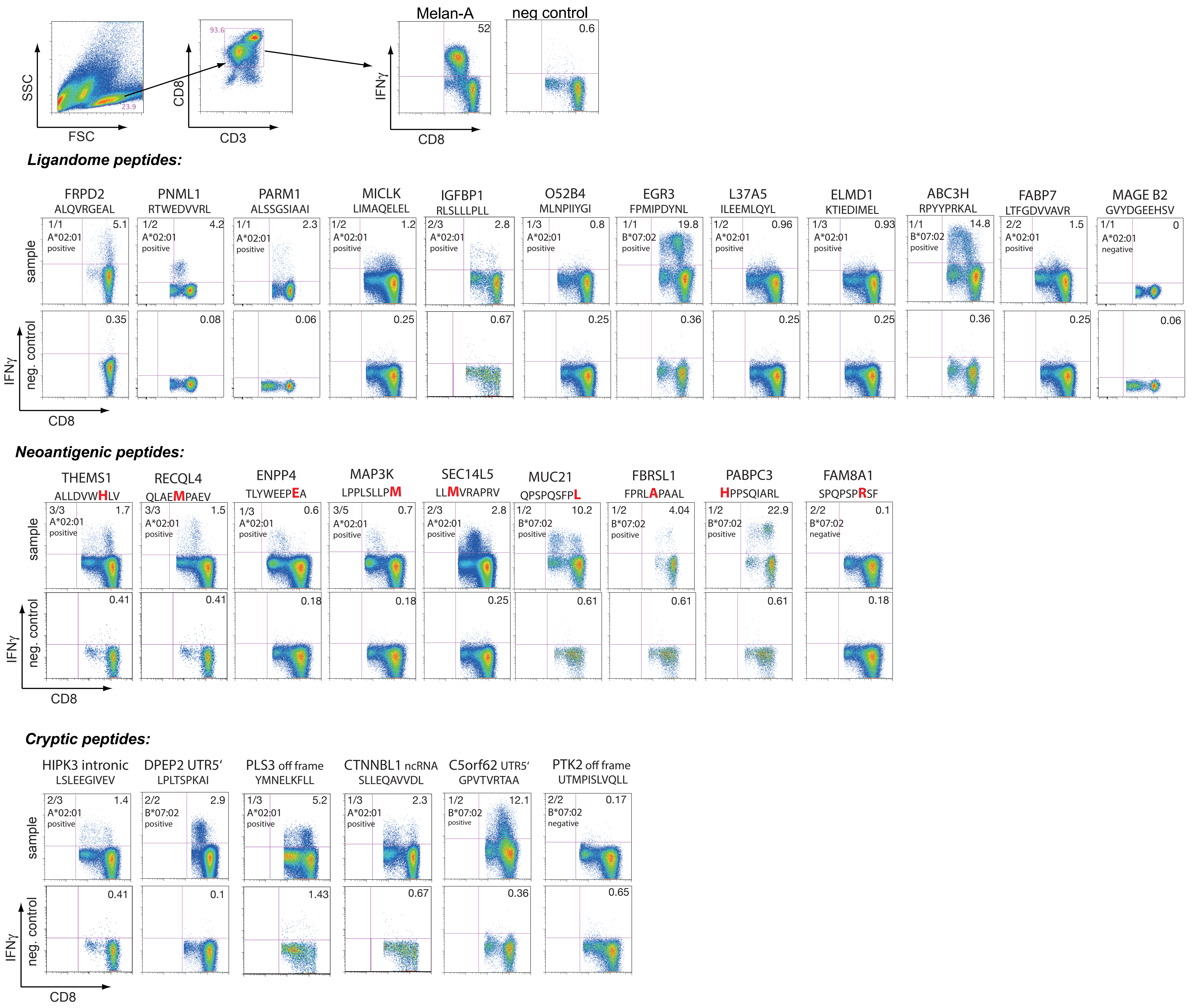


**Supplementary Figure 4:**

Complete data set of priming assays for all 27 selected peptides. Highly purified naïve CD8$^+$ T cells from 5 healthy donors (four HLA-A*02:01, one B07*02) were cocultured for 12 days with peptide-loaded autologous DCs in the presence of IL-7, -15, and -21, restimulated and assayed for IFNγ/TNFα production. A response 3x above negative control (empty DCs) was considered positive, responses above background but but less than 3x were defined as intermediate, responses below the background as negative. Gating strategy is demonstrated in the upper panel. Numbers in the upper right quadrant display percentages of CD8$^+$IFNγ$^+$, numbers in the upper left quadrant indicate number of positive tests against the number of conducted experiments, below the HLA-restricition is indicated as well as the interpretation of the results as positive or negative. The red colored amino acid in the neoantigenic groups indicates the mutation.
